# A rapid review of the effectiveness of innovations to support patients on elective surgical waiting lists

**DOI:** 10.1101/2022.06.10.22276151

**Authors:** Chukwudi Okolie, Rocio Rodriguez, Alesha Wale, Amy Hookway, Hannah Shaw, Alison Cooper, Ruth Lewis, Rebecca-Jane Law, Micaela Gal, Jane Greenwell, Adrian Edwards

## Abstract

Surgical waiting times have reached a record high, in particular with elective and non-emergency treatments being suspended or delayed during the COVID-19 pandemic. Prolonged waits for surgery can impact negatively on patients who may experience worse health outcomes, poor mental health, disease progression, or even death. Time spent waiting for surgery may be better utilised in preparing patients for surgery. This rapid review sought to identify innovations to support patients on surgical waiting lists to inform policy and strategy to address the elective surgical backlog in Wales.

The review is based on the findings of existing reviews with priority given to robust evidence synthesis using minimum standards (systematic search, study selection, quality assessment, and appropriate synthesis). The search dates for prioritised reviews ranged from 2014-2021.

Forty-eight systematic reviews were included. Most available evidence is derived from orthopaedic surgery reviews which may limit generalisability. The findings show benefits of exercise, education, smoking cessation, and psychological interventions for patients awaiting elective surgery. Policymakers, educators, and clinicians should consider recommending such interventions to be covered in curricula for health professionals.

Further research is required to understand how various patient subgroups respond to preoperative interventions, including those from underserved and minority ethnic groups, more deprived groups and those with lower educational attainments. Further research is also needed on social prescribing or other community-centred approaches.

It is unclear what impact the pandemic (and any associated restrictions) could have on the conduct or effectiveness of these interventions.

**Rapid Review Details:** *Review conducted by:* Public Health Wales

*Review Team:*

▪ Dr Chukwudi Okolie
▪ Rocio Rodriguez
▪ Dr Alesha Wale
▪ Amy Hookway
▪ Hannah Shaw

*Review submitted to the WCEC on:* 1^st^ April 2022

*Stakeholder consultation meeting:* 6^th^ April 2022

*Rapid Review report issued by the WCEC in:* June 2022

*WCEC Team:*

▪ Adrian Edwards, Alison Cooper, Ruth Lewis, Becki Law, Jane Greenwell involved in drafting Topline Summary and editing

*This review should be cited as:* RR00030. Wales COVID-19 Evidence Centre. Rapid review of the effectiveness of innovations to support patients on elective surgical waiting lists. April 2022.

*This report can be downloaded here:* https://healthandcareresearchwales.org/wales-covid-19-evidence-centre-report-library

*Disclaimer:* The views expressed in this publication are those of the authors, not necessarily Health and Care Research Wales. The WCEC and authors of this work declare that they have no conflict of interest.

**TOPLINE SUMMARY:** Our rapid reviews use a variation of the systematic review approach, abbreviating or omitting some components to generate the evidence to inform stakeholders promptly whilst maintaining attention to bias. They follow the methodological recommendations and minimum standards for conducting and reporting rapid reviews, including a structured protocol, systematic search, screening, data extraction, critical appraisal, and evidence synthesis to answer a specific question and identify key research gaps. They take 1-2 months, depending on the breadth and complexity of the research topic/ question(s), extent of the evidence base, and type of analysis required for synthesis.

*Who is this summary for?:* Health Boards and others involved in planning, monitoring, managing waiting lists for surgery.

*Background / Aim of Rapid Review:* **Surgical waiting times** have reached a **record high**, in particular with elective and non-emergency treatments being suspended or delayed during the COVID-19 pandemic. **Prolonged waits for surgery can impact negatively on patients** who may experience worse health outcomes, poor mental health, disease progression, or even death. Time spent waiting for surgery may be better utilised in preparing patients for surgery. This rapid review sought to **identify innovations to support patients on surgical waiting** lists to inform policy and strategy to address the elective surgical backlog in Wales. The **review is based on the findings of existing reviews** with priority given to robust evidence synthesis using minimum standards (systematic search, study selection, quality assessment, and appropriate synthesis).

*Key Findings:* Extent of the evidence base

▪ 48 systematic reviews were included; **17 reviews were prioritised for inclusion in the narrative synthesis**. A further 10 protocols of ongoing systematic reviews were included.
▪ Most reviews (n=23) focused on **orthopaedic surgical procedures**.
▪ Most reviews (n=31) focussed on **exercise-based interventions**. Other interventions were **educational** (n=6), **psychological** (n=2), **smoking cessation** (n=1), **weight loss** (n=1), and **multicomponent interventions** (n=7).
▪ There were **limited data provided on socio-demographic characteristics** of patients.
▪ No review evaluated the impact of the intervention on surgical treatment.
▪ **No evidence** relating to the use of **social prescribing or other community-centred approaches** to support surgical wait-listed patients was identified.
▪ **No evidence** was identified in the context of the **current COVID-19 pandemic**. Recency of the evidence base

▪ The search dates for the prioritised reviews ranged from 2014-2021; these were conducted in 2020 (n=3) or 2021 (n=3) for six reviews. Evidence of effectiveness

▪ **Preoperative exercise interventions** (n=9; 6 were orthopaedic) **could help improve preoperative and postoperative outcomes** such as **pain, muscle strength and function, and reduced incidence of postoperative complications**, in people awaiting elective surgery.
▪ **Educational interventions** (n=3; 1 was orthopaedic) were **effective at improving knowledge** in patients awaiting elective surgery. However, the evidence about these interventions improving pre- and postoperative **pain and physical functioning** in orthopaedic patients **is limited**. There were **mixed findings for the effectiveness** of preoperative educational interventions on **psychological outcomes.**
▪ **Psychological interventions** (n=2; 1 was orthopaedic) **evidence is limited** but indicates it **may have a positive effect on anxiety and mental health components** of quality of life postoperatively. The evidence in support of such interventions in reducing postoperative pain is inconclusive.
▪ **Smoking cessation interventions** (n=1) providing behavioural support and offering nicotine replacement therapy i**ncreased short-term smoking cessation and may reduce postoperative morbidity**. Intensive preoperative smoking cessation interventions appear to reduce the incidence of postoperative complications, but not brief interventions.
▪ **Multicomponent interventions** (n=2; 1 was orthopaedic) consisting of both exercise and education components **could shorten the length of hospital stay and improve postoperative pain, function, and muscle strength**. Best quality evidence
Three reviews were treated as high quality. Two evaluating exercise-based interventions (Fenton et al. 2021; Katsura et al. 2015) and one psychological preparation (Powell et al. 2016).

*Policy Implications:*

▪ Most available evidence is derived from **orthopaedic surgery** reviews which may **limit generalisability**.
▪ These findings **show benefits of exercise, education, smoking cessation, and psychological interventions for patients awaiting elective surgery**. Policymakers, educators and clinicians should consider recommending **such interventions to be covered in health professionals’ curricula**.
▪ Further research is required to understand how various **patient subgroups** respond to preoperative interventions, including those from **underserved and minority ethnic groups, more deprived groups and those with lower educational attainments**.
▪ Further research is needed on **social prescribing or other community-centred approaches**.
▪ It is unclear what impact the pandemic (and any associated restrictions) could have on the conduct or effectiveness of these interventions.

*Strength of Evidence:* The primary studies included in the reviews were mainly randomised controlled trials, but most had small sample size, varied by surgical type, and often had issues regarding blinding.

## 1. BACKGROUND

This Rapid Review is being conducted as part of the Wales COVID-19 Evidence Centre Work Programme. The above question was suggested by Cwm Taf UHB, All Wales Medical Directors, Royal College of Surgeons Edinburgh, and the TAG modelling subgroup.

### 1.1 Purpose of this review

The COVID-19 pandemic has stretched hospital resources and led to huge waiting lists for elective surgical treatment in Wales and globally. This, in addition to the significant elective surgery backlog that existed prior to the pandemic, has resulted in a massive number of vulnerable patients waiting for surgical procedures. Prolonged waits for surgery can impact negatively on patients who may experience worse health outcomes, disease progression, or even death. Patients on waiting lists for time-sensitive surgeries may also experience severe psychological distress and worse mental health outcomes including increased anxiety and depression (Gagliardi et al., 2021).

As restrictions ease across the UK, various strategies are being introduced to address the growing elective surgical patient backlog. These include increasing surgical activity by implementing ‘demand-side’ interventions such as prioritisation of cases and pooling of waiting lists (Carr et al., 2021), and ‘supply-side’ interventions such as establishment of ‘green’ or COVID-light sites (Royal College of Surgeons of England, 2021), and enhancing adequate hospital and workforce capacity (Royal College of Surgeons of England, 2020). However, there have been calls for an NHS-wide commitment to transform not just ‘how long’ patients wait, but ‘how’ patients wait (Centre for Perioperative Care, 2021). It is now acknowledged that the time spent waiting for surgery can be better utilised in preparing patients for surgery. Turning the ‘waiting list’ into ‘preparation lists’ will allow patients to be fully supported to use the waiting period proactively, and is expected to improve postoperative clinician and patient reported outcomes, help reduce late cancellations, as well as reduce the length of time patients stay in hospital (Levy et al., 2021).

The purpose of this rapid review is to identify innovations to support patients on surgical waiting lists so as to inform strategy and policy to address the elective surgical backlog in Wales. To address this we looked to answer the question ‘What are the effectiveness of innovations to support patients on elective surgical waiting lists?’

## 2. RESULTS

### 2.1 Overview of the Evidence Base

A total of 58 secondary sources (48 full systematic reviews and 10 systematic review protocols) were identified for inclusion in this rapid review based on our eligibility criteria (section 5.2). The interventions and outcomes covered in the 48 published systematic reviews have been mapped in Table 1. A summary of the 10 ongoing systematic reviews can be found in appendix 4.

**Table 1:**
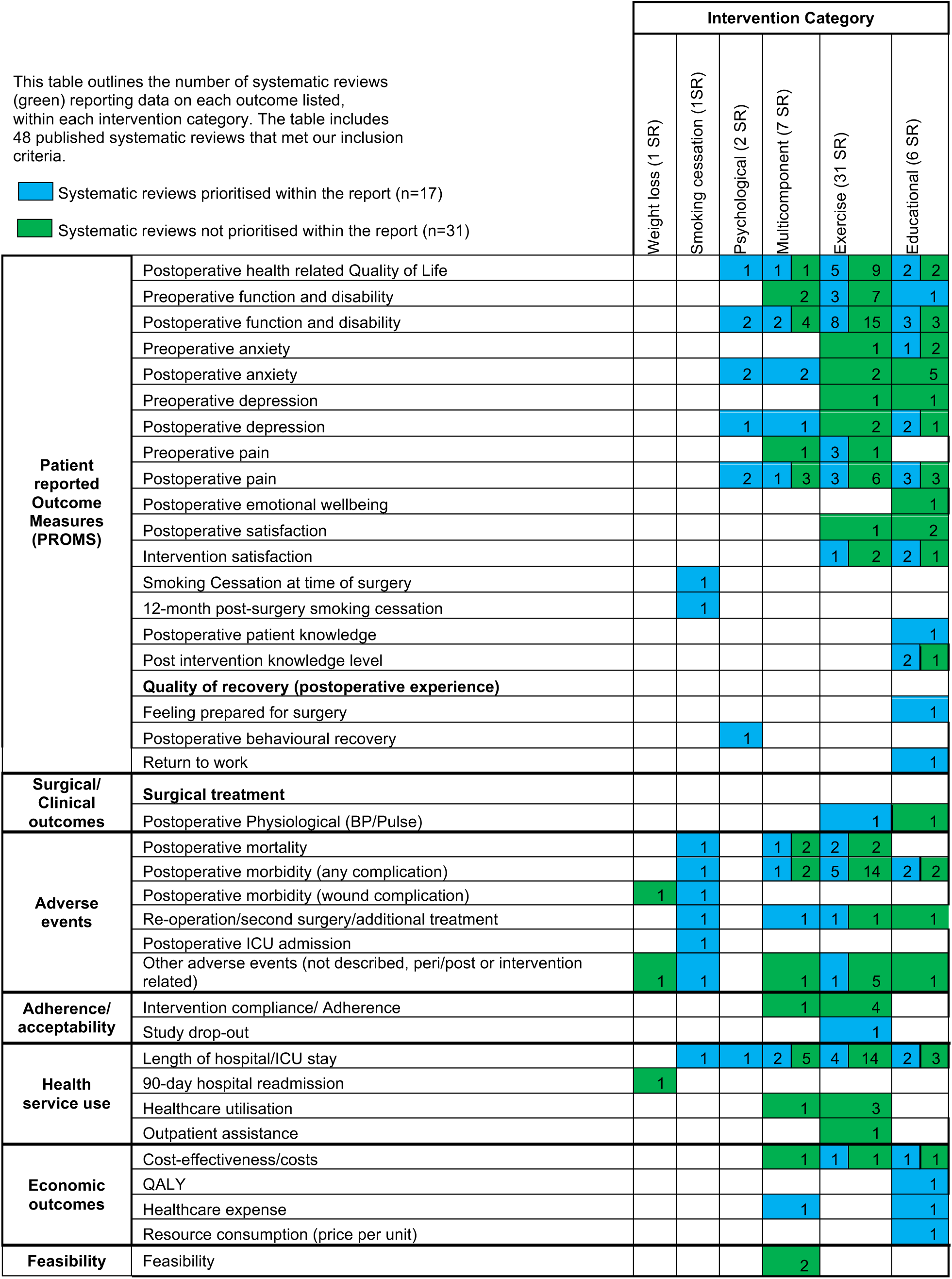
Outcomes mapped by intervention category

Of the **48 published systematic reviews**, the majority focussed on orthopaedic surgical procedures (n = 23). Other systematic reviews focussed on cardiac surgery (n = 7), vascular surgery (n = 4), cardiac and abdominal surgery (n = 2), cardiac, abdominal and orthopaedic surgery (n = 2), and cardiac and vascular surgery (n = 1). Nine systematic reviews did not have a specific surgical focus. The majority of systematic reviews focused on exercise-based interventions (n = 31). Other interventions were educational (n = 6), psychological (n = 2), smoking cessation (n = 1), and weight loss interventions (n = 1). Seven reviews focused on multicomponent interventions.

In light of the large number of secondary sources identified and the timeframe required to conduct this rapid review, we undertook a prioritisation process (outlined in detail in section 5), allowing us to scrutinise in more detail a smaller number of systematic reviews. After applying the prioritisation process, we included 17 systematic reviews for critical appraisal, and synthesis. A detailed summary of the prioritised systematic reviews can be found in Table 2. A summary of the data extraction for the remaining 31 systematic reviews that met our inclusion criteria, but were not included in the focussed synthesis, along with the reasons why they were not prioritised are outlined in appendix 3. **From this point onwards, we refer only to the 17 prioritised systematic reviews**.

**Table 2:** Summary of included reviews

| Citation | Review details | Included studies | Quality* | Findings and observations/notes |
| --- | --- | --- | --- | --- |
| **Exercise-based interventions** |  |  |  |  |
| Alshewaier et al. (2017) <u>The effectiveness of pre-operative exercise physiotherapy rehabilitation on the outcomes of treatment following anterior cruciate ligament injury: a systematic review</u>. Clinical rehabilitation, 31(1), pp.34-44. | **Intervention Type:** Preoperative exercise physiotherapy rehabilitation (prehabilitation)<br>**Review period:** Inception of databases to December 2015<br>**Review purpose:** To evaluate the effectiveness of preoperative exercise physiotherapy rehabilitation on the outcomes of treatment following anterior cruciate ligament injury.<br>**Included study designs:** RCTs<br>**Included outcome measures:** Pain, quality of life, physical knee function, swelling, range of motion, muscle strength and functional activity. | **Number of included studies:** 8<br>**Key characteristics:** The eight studies investigated a total of 451 subjects, of which 71% (n = 319) were male participants. The age of the participants ranged from 15 to 57 years. Preoperative rehabilitation protocols were different in their content, duration and frequency of intervention. The average duration of the preoperative intervention was 14 weeks (range 3-24 weeks). The content of the preoperative intervention consisted of: quadriceps and/or hamstring strengthening exercises; proprioception and/or balance training; gait re-education; treatment to increase range of motion; functional specific rehabilitation and plyometrics. The primary studies originated from Sweden (n = 4), USA (n=2), Ireland (n = 1), **UK (n = 1)** | Low | **Pain** was used as an outcome in three studies. No significant difference was found in patient reported pain between the intervention and control groups in any of the studies.<br>**Physical function** (in recreational or sports activities) was used as an outcome in seven of the eight studies. Two studies found a significant improvement in physical function in the intervention group compared to the control.<br>One study found a significant increase in function from baseline to preoperatively and at 12 weeks postoperatively, in the intervention group who received preoperative physiotherapy rehabilitation compared to the control group which received no preoperative physiotherapy intervention. Five studies found no significant difference in physical function between the groups.<br>**Quality of life** was examined in three studies using a subscale of the Knee Injury and Osteoarthritis Outcome Score. Whilst there was a significant improvement in quality of life from baseline following intervention in both groups, none of the studies reported any significant difference in quality of life between the control and intervention groups.<br>**Range of motion** was used as an outcome in only one study. There was no significant difference in range of motion between the two rehabilitation programmes using open and closed kinetic chain exercises. |
|  |  |  |  | **Muscle (quadriceps and/or hamstring) strength and function** were measured in four studies. One study reported that the intervention group had greater quadriceps muscle strength, however, no other significant differences in strength were found.<br>One study found that quadriceps strength increased in both groups, although there was no significant difference between the groups. However, they did find that quadriceps strength and knee excursions were more symmetrical 6 months postoperatively in the intervention group that received perturbation training and progressive quadriceps strength training than the control group who received strength training alone. The remaining two studies found no significant difference in muscle strength between the intervention and control groups.<br>The outcomes of **knee-related symptoms, including swelling**, were measured in four studies. No significant differences in symptoms between the control and intervention groups were found. |
| Blasco et al. (2021)<br>Sensorimotor training prior total knee arthroplasty and effects on functional outcome: A systematic review and meta-analysis.<br>Gait & Posture, 86, pp.83-93. | **Intervention Type:** Preoperative sensorimotor training<br>**Review period:** Up to May 3, 2020<br>**Review purpose:** To determine whether preoperative sensorimotor training is effective in improving the functional outcome in patients undergoing total knee arthroplasty.<br>**Included study designs:** RCTs<br>**Included outcome measures:** Functional outcome, balance, pain, knee function, quality of life, and falls | **Number of included studies:** 5<br>**Key characteristics:** Overall, 332 participants with a mean age of 69.9 (65.1–72.8) years were randomized. All study participants were awaiting total knee arthroplasty, presenting with severe knee osteoarthritis. Countries of included studies were not described. | Low | **Preoperative effects of sensorimotor training**<br>There was limited evidence suggesting that preoperative sensorimotor training enhanced self-reported function (SMD, 0.89; 95 % CI, 0.16–1.62), functional performance (SMD, 0.56; 95 % CI, 0.19 to 0.93), or knee function (SMD = 0.22–1.05) compared with conventional care.<br>Sensorimotor training produced greater preoperative benefits than standard care on dynamic (SMD, 1.61; 95 % CI, -0.10 to 3.31) and overall state of balance (SMD, 0.84; 95 % CI, 0.36–1.31) and was more effective in reducing pain (SMD, 0.68; 95 % CI, 0.09–1.26).<br>**Postoperative effects**<br>Overall, there was high quality evidence that the superior functional preoperative benefits registered in<br>participants implementing sensorimotor training were only maintained in terms of functional performance till 6 weeks after surgery.<br>The training programme was no more effective than standard care in enhancing balance or resolving pain. Likewise, the quality of life of all the included participants was similar one year after surgery, regardless of groups and interventions (SMD, -0.14; 95 % CI, -0.62 to 0.34).<br>Moderate quality evidence suggested that benefits were only maintained in terms of functional performance up to 3 months after surgery (SMD = 0.37; 95 % CI, 0.13 to 0.62).<br>The outcome was similar after one year, regardless of whether such training is implemented. |
| Fenton et al. (2021)<br><u>Prehabilitation exercise therapy before elective abdominal aortic aneurysm repair</u>.<br>Cochrane Database of Systematic Reviews 2021, Issue 7. Art. No.: CD013662.<br>DOI: 10.1002/14651858.CD013662.pub2. | **Intervention type:** Prehabilitation exercise<br>**Review period:** Inception of databases to July 2020<br>**Review purpose:** To assess the effects of exercise programmes on perioperative and postoperative morbidity and mortality associated with elective abdominal aortic aneurysm (AAA) repair<br>**Included study designs:** RCTs<br>**Included outcome measures:**<br>Primary outcomes<br>• 30-day (or longer if reported) mortality post-AAA repair<br>• Perioperative and postoperative complications | **Number of included studies:** 4<br>**Key characteristics:** Four trials with a total of 232 participants were included in the review. All four trials compared exercise versus usual care, but one trial did not describe the components of the usual care implemented. Exercise regimens implemented in the included trials varied, although most studies implemented at least two sessions weekly for a minimum of two weeks prior to surgery. The primary studies originated from the **UK (n = 3)**, and the Netherlands (n = 1). | High | **Primary outcomes**<br>**30-day (or longer if reported) mortality post-AAA repair**<br>Overall, it is uncertain whether prehabilitation exercise reduces the occurrence of 30-day (or longer if reported) mortality post-AAA repair (RR 1.33, 95% CI 0.31 to 5.77; 3 trials, 192 participants; very low-certainty evidence). There was no statistical heterogeneity between studies ($I^2 = 0\%$, $P = 0.55$).<br>**Perioperative and postoperative complications: cardiac complications**<br>Data from one trial with 124 participants showed that overall, prehabilitation exercise may decrease the occurrence of cardiac complications compared to usual care (RR 0.36, 95% CI 0.14 to 0.92; 1 trial, 124 participants; low-certainty evidence).<br>**Perioperative and postoperative complications: pulmonary complications**<br>Overall, it is uncertain whether prehabilitation exercise decreases the occurrence of pulmonary complications |
|  | Secondary outcomes<br>• Length of intensive care unit (ICU) stay<br>• Length of hospital stay<br>• Number of days on a ventilator<br>• Change in aneurysm size pre- and post-exercise<br>• Quality of life (QoL) |  |  | compared to usual care (RR 0.49, 95% 0.26 to 0.92; 2 trials, 144 participants; very low certainty evidence). Moderate statistical heterogeneity ($I^2 = 15\%$, $P = 0.31$) was detected.<br>**Perioperative and postoperative complications: renal complications**<br>Overall, prehabilitation exercise may reduce the risk of the occurrence of renal complications compared to usual care (RR 0.31, 95% CI 0.11 to 0.88; 1 trial, 124 participants; low-certainty evidence).<br>**Perioperative and postoperative complications: need for reintervention**<br>It is uncertain whether prehabilitation exercise reduces the need for re-intervention compared to usual care (RR 1.29, 95% 0.33 to 4.96; 2 trials, 144 participants; very low-certainty evidence).<br>**Perioperative and postoperative complications: postoperative bleeding**<br>Overall, it is uncertain whether prehabilitation exercises reduces the occurrence of postoperative bleeding compared to usual care (RR 0.57, 95% CI 0.18 to 1.80; 1 trial, 124 participants; very low certainty evidence).<br>**Secondary outcomes**<br>There was little or no difference between the exercise and usual care (no exercise) groups in length of ICU stay, length of hospital stay and quality of life. None of the studies reported data for the number of days on a ventilator and change in aneurysm size pre- and post-exercise outcomes<br>**Notes**<br>The GRADE assessments showed that the evidence ranged from very low certainty to low certainty. |
| Husted et al. (2020) The relationship between prescribed pre-operative knee-extensor exercise dosage and effect on knee-strength prior to and following total knee arthroplasty: a systematic review and meta-regression analysis of randomized controlled trials. Osteoarthritis and Cartilage, 28(11), pp.1412-1426. | **Intervention Type:** Exercise intervention (resistance training)<br>**Review period:** Up to 27 August 2019<br>**Review purpose:** To evaluate the relationship between prescribed knee-extensor strength exercise dosage in preoperative exercise intervention and the effect on knee-extensor muscle strength prior to and following total knee arthroplasty (TKA).<br>**Included study designs:** RCTs<br>**Included outcome measures:**<br>Primary outcome;<br>Knee-extensor strength<br>Secondary outcomes;<br>Knee pain<br>Patient reported physical function<br>Knee-related performance-based function<br>Adverse events | **Number of included studies:** 12<br>**Key characteristics:** Data were extracted for 616 patients scheduled for TKA (median (range), age 65.9 (62.1–70.7), BMI 31.0 (27.9–34.8), 58.6% female). The prehabilitation interventions included on average 1.8 knee-extensor exercises (range 1–3), an average duration of 6 weeks (range 4–8.9), an average of 4.4 sessions/week (range 2–14), 2.5 knee-extensor sets/session (range 1.5–3.5) and 11 repetitions/set (range 8–15). Countries of included studies were not described | Low | Meta-regression analysis showed no relationship between prescribed preoperative knee-extensor exercise dosage and change in knee-extensor strength neither prior to (slope 0.0005 [95%CI -0.007 to 0.008]) or 3 months following TKA (slope 0.0014 [95%CI -0.006 to 0.009]).<br>No relationship was found between prescribed knee-extensor dosage and knee pain or physical function neither prior to nor 3 months following TKA<br>**Preoperative effects of prehabilitation**<br>Prior to TKA, a moderate effect favoring preoperative exercise for increase in knee-extensor strength was found (SMD 0.50 [95%CI 0.12 to 0.88]), but not at 3 months following TKA (SMD -0.01 [95%CI -0.45 to 0.43]).<br>**Postoperative effects of prehabilitation**<br>Overall, no effect of prehabilitation on knee-extensor strength 3 months following TKA (SMD -0.01 [95% CI -0.45 to 0.43]) was found. No effect of prehabilitation was seen in any postoperative outcomes |
| Katsura et al. (2015) Preoperative inspiratory muscle training for postoperative pulmonary complications | **Intervention type:** Exercise (inspiratory muscle training)<br>**Review period:** up to October 2014<br>**Review purpose:** To assess the effectiveness of preoperative inspiratory muscle training (IMT) on postoperative pulmonary | **Number of included studies:** 12<br>**Key characteristics:** All included trials were single-institution RCTs<br>In the majority of trials, sample sizes per arm were small. Seven of the 12 trials recruited fewer than 20 participants. Three | High | **Primary outcomes**<br>**Postoperative pulmonary complications (PPCs)**<br>**PPCs; atelectasis**<br>Data from seven trials involving 443 participants showed preoperative IMT was associated with a |
| <u>in adults undergoing cardiac and major abdominal surgery</u>.<br>Cochrane Database of Systematic Reviews 2015, Issue 10. Art. No.: CD010356. DOI: 10.1002/14651858.CD010356.pub2. | complications (PPCs) in adults undergoing cardiac or major abdominal surgery<br>**Included study designs:** RCTs<br>**Included outcome measures:**<br>Primary outcomes<br>1. PPCs<br>2. All-cause mortality within 30 days during postoperative period.<br>3. Adverse events from IMT.<br>Secondary outcomes<br>1. Maximal inspiratory muscle strength and endurance<br>2. Duration of hospital stay.<br>3. Other types of complications within 30 days during postoperative period<br>4. Total drop-out from the study<br>5. Quality of life<br>6. Cost analysis. | trials recruited 20 to 40 participants per arm and one trial recruited less than 10 participants per arm. Only one trial recruited over 100 participants to each arm. Overall, 12 trials involving 695 participants were available for assessing the effects of interventions. Of the 12 trials, five were performed in the Netherlands; five in Brazil; one in the United Kingdom; and one in Israel. Five trials included participants awaiting elective cardiac surgery; the remaining seven trials included participants awaiting elective major abdominal surgery. Three trials did not report the number of female participants. One trial recruited only women. In the remaining eight trials, the proportion of females ranged between 22% and 78%. Ten of the 12 included trials provided the mean (or median) age of participants. One trial recruited young participants, whose mean age was 35 years. Seven trials recruited middle-age participants, whose mean age was in the 50s to 60s. The remaining two trials recruited elderly individuals only, whose mean age was in the 70s. Primary studies originated from the Netherlands (n = 5), Brazil (n = 5), the UK (n = 1) and Israel (n = 1) |  | reduction of postoperative atelectasis, compared with usual care or non-exercise intervention (RR 0.53; 95% CI 0.34 to 0.82; P value = 0.004; I2 statistic = 0%).<br>**PPCs; pneumonia**<br>Data from 11 trials involving 675 participants showed preoperative IMT was associated with a reduction of postoperative pneumonia, compared with usual care or no exercise intervention (RR 0.45; 95% CI 0.26 to 0.77; P value = 0.004; I2 statistic = 0%)<br>**PPCs; mechanical ventilation exceeding 48 hours**<br>Four trials reported on postoperative length of mechanical ventilation. Pooled analysis showed there was no evidence of a reduction of postoperative mechanical ventilation exceeding 48 hours in the groups receiving preoperative IMT (RR 0.55; 95% CI 0.03 to 9.20; P value = 0.68; I2 statistic = 56%)<br>**All-cause mortality within 30 days during postoperative period**<br>Seven trials reported on all-cause mortality within the postoperative period. In four trials, there were zero events in both groups. Data from the remaining three trials, were pooled in a meta-analysis which found the effect of IMT on all-cause postoperative death is uncertain (RR 0.40; 95% CI 0.04 to 4.23; P value = 0.09; I2 statistic = 59%).<br>**Adverse events**<br>Eight trials reporting on adverse events reported that there were none in both study groups<br>**Secondary outcomes**<br>**Maximal inspiratory muscle strength; Pi-max**<br>Analysis of three trials found that preoperative IMT was not associated with improved Pi-max (mean<br>difference (MD) -7.87; 95% CI -21.36 to 5.61; P value = 0.25; I2 statistic = 0%)<br>**Duration of hospital stay**<br>The analysis of six trials found that preoperative IMT was associated with reduced length of hospital stay (MD -1.33; 95% CI -2.53 to -0.13; P value = 0.03; I2 statistic = 7%).<br>**Other types of complications within 30 days during postoperative period**<br>One trial reported the occurrence of other types of early postoperative complications: cardiac complications, neurological complications, or surgical-site infections. This trial reported that there is no statistically significant difference regarding postoperative heart failure; experimental 1/15 (67%) versus control 1/15 (67%), P value = 0.65.<br>**Total drop-out from the study for any reason, as surrogate measure of overall acceptability**<br>Ten trials reported on dropouts, however five trials reported zero events, therefore post-hoc analysis conducted found no evidence of significant difference between groups (RD -0.00; 95% CI -0.01 to 0.01; P value = 0.95; I2 statistic = 0%).<br>**Quality of life**<br>One trial with 42 participants reported on quality of life. In this trial, there were no significant differences across the groups as measured by the European Organisation for Research and Treatment of Cancer Quality of Life Questionnaire<br>No trials reported on cost analysis.<br>**Notes**<br>Review authors used the GRADE approach to assess the quality of the body of evidence associated with<br>specific review outcomes. The overall quality of studies for the incidence of pneumonia was moderate, whereas the overall quality of studies for the incidence of atelectasis, all-cause postoperative death, adverse events, and duration of hospital stay was low or very low.<br>Review authors conducted funnel plot analyses for the incidence of pneumonia. The funnel plot showed asymmetrical appearance with a gap in the bottom corner of the graph, but Egger's test was not statistically significant (P value = 0.22). When we carried out the trim-and-fill imputation as a sensitivity check, the overall results did not change materially (the imputed RR = 0.41). |
| Potts et al. (2021) The effectiveness of preoperative exercise programmes on quadriceps strength prior to and following anterior cruciate ligament (ACL) reconstruction: A systematic review. Physical Therapy in Sport. 2022-03-01, Volume 54, Pages 16-28 | **Intervention Type:** Preoperative strength based exercise<br>**Review period:** Inception of databases to March 2021<br>**Review purpose:** To evaluate the effectiveness of preoperative exercise programmes on quadriceps strength prior to and following anterior cruciate ligament (ACL) reconstruction.<br>**Included study designs:** RCTs, prospective cohort studies<br>**Included outcome measures:** Quadriceps strength | **Number of included studies:** 10<br>**Key characteristics:** Six of the included studies were RCTs and four were prospective cohort studies. There was a total of 457 participants across all studies (303 male, 134 female, 20 undisclosed). Participant age ranged from 15 to 57 years. The frequency and duration of preoperative exercise ranged from once to twice daily home exercise over 6 weeks, 10 supervised sessions within a maximum of 5-weeks, and up to three sessions weekly over 16 weeks resulting in a mean (13–63) 34 sessions. The studies originated from USA (n = 4), Australia (n = 2), and had one study each from Ireland, India, Korea and Sweden | Low | Five studies demonstrated preoperative exercise of 4–16 weeks duration can significantly increase preoperative quadriceps strength. One study demonstrated preoperative open kinetic chain (OKC) exercise produced significantly stronger preoperative quadriceps compared to closed kinetic chain (CKC) exercise. One study showed no between group (intervention vs control) quadriceps strength difference pre or 12 weeks postoperatively.<br>Due to heterogeneity, participant numbers and with only 6 of the 10 studies reporting quadriceps strength increases after an exercise intervention the level of evidence is categorised as 'limited' |
| Rodrigues et al. (2021) <u>Effectiveness of preoperative breathing exercise interventions in patients undergoing cardiac surgery: A systematic review</u>. Revista Portuguesa de Cardiologia (English Edition), 40(3), pp.229-244. | **Intervention type:** Exercise (inspiratory muscle training)<br>**Review period:** Inception of databases to October 2019<br>**Review purpose:** To discover whether breathing therapy (any breathing exercise) performed preoperatively in persons awaiting cardiac surgery is effective, when comparing the following postoperative outcomes: respiratory parameters, postoperative pulmonary complications (PPC) and length of hospital stay (LOS), among cardiac surgery patients included in a preoperative breathing therapy program and those who were not included in any program preoperatively.<br>**Included study designs:** Randomised controlled trials (RCT) and cohort studies<br>**Included outcome measures:** Postoperative respiratory parameters, length of hospital stay and pulmonary complications | **Number of included studies:** 11<br>**Key characteristics:** Ten RCTs and one prospective cohort study (representing 1240 participants) were included. The median sample size of the 11 selected studies was 70 [range: 26-346]. Median age of all patients included was 62 years old [range: 54-71]. Male gender accounted for the main subjects of each trial with a median percentage of 69% [range: 50%-100%], whereas female gender presented a median percentage study size of 31% [range: 0%-50%].<br>All studies aimed to improve the quality of respiratory performance after cardiac surgery. Countries of included studies were not described | Critically low | **Postoperative pulmonary complications**<br>Based on data from eight trials (1077 participants), there was a significant reduction in the relative risk of developing PPC with preoperative breathing therapy exercises. When results from trials included in this meta-analysis were pooled, there was moderate heterogeneity, and the pooled risk of developing PPC was 0.47 (CI95% 0.26 to 0.85).<br>A subgroup analysis assessing the benefits of the preoperative intervention in the elderly ($\geq 65$ years old) reported a significant reduction in the risk of developing PPC 0.30 (CI 95% 0.19 to 0.49).<br>**Length of hospital stay**<br>Data from seven trials (1050 participants) showed a reduction in LOS in the intervention group with a pooled mean difference of 0.81 days (CI 95% 0.48 to 1.38), however this was not statistically significant.<br>**Postoperative respiratory improvement**<br>Preoperative breathing exercise interventions helped improve postoperative ventilation in the analyses of four trials (219 participants), a pooled mean difference of -25.36 (CI 95% -31.87 to -18.85) with substantial heterogeneity.<br>**Notes**<br>A funnel plot of all 11 studies, was used to check publication bias; minimal asymmetry indicates lack of publication bias |
| Wang et al. (2016) <u>Does preoperative rehabilitation for patients planning to undergo joint</u> | **Intervention Type:** Preoperative rehabilitation programmes (i.e., prescribed and supervised exercises or physiotherapy with or without cointerventions such as education, nutritional counselling, acupuncture, transcutaneous | **Number of included studies:** 22<br>**Key characteristics:** In total, 22 RCTs (1492 patients) of prehabilitation versus no prehabilitation met the inclusion criteria. Most studies were conducted | Low | **Postoperative pain**<br>Fifteen trials with 18 comparisons and 1046 patients reported postoperative pain scores using different instruments. Prehabilitation significantly reduced postoperative pain at 4 weeks or less; however, the reduction of pain was clinically nominal (4 trials, 213 patients, WMD -6.1 points, 95% CI -10.6 to -1.6 points, |
| replacement surgery improve outcomes? A systematic review and meta-analysis of randomised controlled trials. BMJ open, 6(2), p.e009857. | electrical nerve stimulation, etc.)<br>**Review period:** Up to 10 November 2015<br>**Review purpose:** To assess the clinical impact of prehabilitation before joint replacement.<br>**Included study designs:** RCTs<br>**Included outcome measures:** Postoperative pain scores (Visual Analogue Scale (VAS), or pain subcomponents of Western Ontario and McMaster Universities osteoarthritis index (WOMAC) or pain-related subdomains of other instruments), Patient functionality (WOMAC function score, SF-36 physical functioning subdomain or other function-related instruments), time to resume activities of daily living (ADL), quality of life, patient satisfaction, infection, transfusions, stroke and death or overall postoperative complications, hospital length of stay, readmissions and total hospital costs or total health system costs. | in North America and Europe. The median sample size of included studies was 54, ranging from 21 to 165 patients. Mean age ranged from 51 to 76 years. The primary studies originated from the USA (n = 4), Australia (n = 3), **UK (n = 2)**, and had two studies each from Canada, Italy, the Netherlands and one study was conducted in each of: Denmark, Thailand, Turkey, Sweden, Switzerland, Greece and Serbia |  | on a scale of 0–100). GRADE: low certainty in estimates.<br>Differences in WOMAC pain scores after 4 weeks were no longer statistically significant for prehabilitation versus control (WOMAC pain score at 6–8 weeks, 5 trials, 488 patients, WMD –1.4, 95% CI –5.5 to +2.6; at 12 weeks, 10 trials, 806 patients, WMD –2.9, 95% CI –6.2 to +0.3; at 24 weeks, 3 trials, 247 patients, –2.5, 95% CI –5.6 to +0.6; at 1 year, 1 trial, 109 patients, WMD –2.0, 95% CI –7.5 to +3.5; GRADE: low to moderate certainty in estimates.<br>**Postoperative function**<br>Prehabilitation slightly improved WOMAC function score at 6–8 (5 trials, 488 patients, WMD –3.9, 95% CI –7.6 to –0.3, RR=1.10; GRADE: moderate certainty in estimates) and 12 weeks (12 trials, 836 patients, WMD –4.0, 95% CI –7.5 to –0.5, RR=1.02, GRADE: very low certainty in estimates). No significant difference for WOMAC function score was found after 12 weeks.<br>**Resumption of activities of daily living (ADL)**<br>Prehabilitation slightly improved time to climbing stairs (2 trials, 99 patients, WMD –1.4 days, 95% CI –1.9 to –0.8 days), toilet use (2 trials, 99 patients, –0.9 days, 95% CI –1.3 to –0.5 days) and chair use (2 trials, 99 patients, WMD –1.2 days, 95% CI –1.7 to –0.8 days), but not time to first day of walking (2 trials, 99 patients, –0.2-day, 95% CI –0.4 to +0.0-day).<br>Effects were similar for knee and hip surgery. Differences were not found for SF-36 scores, length of hospital stay and total cost. Other outcomes of interest were inadequately reported. |
| Wang et al. (2021) A systematic | **Intervention Type:** Preoperative exercise | **Number of included studies:** 12 | Low | **Effect of preoperative exercise intervention on postoperative knee ROM** |
| <u>review and meta-analysis of the effect of preoperative exercise intervention on rehabilitation after total knee arthroplasty.</u><br>Annals of palliative medicine, 10(10), pp.10986-10996. | **Review period:** January 2000 to January 2021<br>**Review purpose:** To investigate the effect of preoperative exercise intervention on rehabilitation before total knee arthroplasty (TKA) by literature retrieval and meta-analysis.<br>**Included study designs:** RCTs<br>**Included outcome measures:**<br>(I) Range of Motion, ROM; (II) The Western Ontario and McMaster Universities Osteoarthritis Index (WOMAC) (6);<br>(III) Visual Analogue Scale (VAS);<br>(IV) Timed Up and Go (TUG) test; (V) 6-Minute Walk Test (6 MWT); (VI) Stair Climbing Test (SCT); (VII) quadriceps strength; (VIII) The Short Form of Health Survey Questionnaire (SF-36); (IX) the Knee Injury and Osteoarthritis Outcome Score (KOOS), and (X) the Berg Balance Scale (BBS). | **Key characteristics:** Twelve studies comprising 889 patients were included in this review. All studies included in the meta-analysis reported the postoperative observation time points at 3, 6, and 12 weeks. Countries of included studies were not described |  | Data from seven studies showed that there was significant difference in ROM flexion in the experimental group compared to controls (MD =4.28, 95% CI: 2.28 to 6.28; Z=4.19; P<0.0001).<br>**Effect of preoperative exercise intervention on quadriceps strength after surgery**<br>Meta-analysis combining data from four studies showed that there was significant difference in quadriceps strength values between the experimental group and control group (MD =1.86; 95% CI: 0.58–3.15; Z=2.84; P=0.005)<br>**Effect of preoperative exercise intervention on postoperative arthritis index score (WOMAC)**<br>Meta-analysis combining data from seven studies showed that there was significant difference in WOMAC score between the experimental group and control group (MD =-10.59; 95% CI: -11.88 to -9.29; Z=16.03; P<0.00001)<br>**Effect of preoperative exercise intervention on postoperative standing and walking test (TUG) score**<br>Data from five studies showed that there was significant difference in TUG score between the experimental group and the control group (MD =-1.29; 95% CI: -1.90 to -0.67; Z=4.08; P<0.0001)<br>**Effect of preoperative exercise intervention on postoperative SF-36 score**<br>Data from five studies showed a significant difference in the quality of life score values between the experimental group and the control group (MD =1.66; 95% CI: 1.13–2.20; Z=6.08; P<0.00001)<br>**Notes**<br>Sensitivity analysis was not performed because statistical heterogeneity was not found between the studies. The funnel plot of the ROM index with the<br>highest number of included articles showed that the two groups of funnels were unevenly distributed on both sides, suggesting a possible publication bias. |
| Smoking cessation |  |  |  |  |
| Citation | Review details | Included studies | Quality | Findings and observations/notes |
| Thomsen et al. (2014)<br>Interventions for preoperative smoking cessation.<br>Cochrane Database of Systematic Reviews. Issue 3. Art. No.: CD002294.<br>DOI: 10.1002/14651858.CD002294.pub4. | **Intervention Type:** Smoking cessation<br>**Review period:** up to January 2014<br>**Review purpose:** To assess the effect of preoperative smoking interventions on smoking cessation at the time of surgery and 12 months postoperatively, and on the incidence of postoperative complications.<br>**Included study designs:** RCTs<br>**Included outcome measures:** Smoking cessation: Prevalence of smoking cessation at the time of surgery, and 12 months postoperatively.<br>Morbidity and mortality: Wound-related complications; secondary surgery; cardiopulmonary complications; admission to intensive care; intra and postoperative mortality; length of stay | **Number of included studies:** 13<br>**Key characteristics:** Thirteen trials representing 2,010 participants were included. Trials were conducted in Denmark, Australia, Canada, the USA, the UK and Sweden between 2002 and 2013. Eleven trials evaluated behavioural interventions, with pharmacotherapy offered in some. Two trials evaluated pharmacotherapy alone. The primary studies originated from Denmark (n = 4), USA (n = 3), Canada (n = 3) and had **one study each from the UK**, Australia and Sweden | Low | **Smoking cessation at the time of surgery**<br>Of the 10 studies evaluating behavioural interventions versus a control, nine reported cessation outcomes. In six studies the intervention achieved a significant increase in smoking cessation at the time of surgery, and one had a lower confidence interval (CI) of 1.<br>Pooling the two trials using intensive interventions gave a RR of 10.76; 95% CI 4.55 to 25.46, and no evidence of heterogeneity. The pooled estimate for the six trials of brief interventions was smaller but also excluded no effect (RR 1.30; 95% CI 1.16 to 1.46. There was still marked heterogeneity ($I^2 = 75\%$). Exclusion of Shi 2013, in which the only difference between groups was measurement of exhaled CO on the morning of surgery, increased the point estimate to RR 1.47; 95% CI 1.27 to 1.70, and reduced $I^2$ to 52%.<br>**Smoking cessation postoperatively at 12-month follow-up**<br>Only five of the 13 studies monitored longer-term postoperative cessation. The two trials of intensive interventions retained significantly higher quit rates in intervention versus control group participants; 23% versus 4%, and 37% versus 17%. The pooled RR was 2.96; 95% CI 1.57 to 5.55 (209 participants) for intensive intervention.<br>Quit rates in the two studies using brief interventions decreased over time and significant differences between intervention and control groups were not maintained at 12 months; 20% versus 20% in Ratner 2004; 12% versus |
|  |  |  |  | 8% in Thomsen 2010, pooled RR 1.09; 95% CI 0.68 to 1.75 (341 participants).<br>**Morbidity and mortality**<br>**Any complication**<br>Two studies, both offering intensive preoperative smoking cessation interventions, found a reduced incidence of postoperative complications. None of the four studies offering brief interventions detected significant differences between intervention and control participants in the incidence of postoperative complications.<br>Pooling intensive and brief interventions separately, the RR for developing any complication was 0.42; 95% CI 0.27 to 0.65 (210 participants) using intensive interventions and 0.92; 95% CI 0.72 to 1.19 (493 participants) for brief interventions. There was no evidence of statistical heterogeneity in either subgroup.<br>**Wound complication**<br>Pooling studies according to intervention intensity, there was an effect of intensive interventions on wound complications: RR 0.31; 95% CI 0.16 to 0.62 (210 participants) but not for brief interventions: RR 0.99; 95% CI 0.70 to 1.40 (325 participants)<br>**Mortality**<br>There were two deaths in the control group during the perioperative period in Sørensen 2003a. In OstroH 2013, one intervention participant and two control participants died postoperatively.<br>**Notes**<br>No studies detected significant differences in duration of hospital admission. No studies reported serious adverse events. No studies detected significant differences between groups in regard to postoperative pulmonary or cardiovascular complications |
|  |  |  |  | No mention of publication bias. Most studies were judged to be at low risk of bias, but the overall quality of evidence was moderate due to the small number of studies contributing to each comparison. |
| Psychological interventions |  |  |  |  |
| Citation | Review details | Included studies | Quality | Findings and observations/notes |
| Powell et al. (2016)<br>Psychological preparation and postoperative outcomes for adults undergoing surgery under general anaesthesia.<br>Cochrane Database of Systematic Reviews, Issue 5. Art. No.: CD008646.<br>DOI: 10.1002/14651858.CD008646.pub2. | **Intervention Type:** Psychological interventions<br>**Review period:** Up to 7 July 2015<br>**Review purpose:** To review the effects of psychological preparation on postoperative outcomes in adults undergoing elective surgery under general anaesthetic.<br>**Included study designs:** RCTs<br>**Included outcome measures:**<br>Primary outcome measures – Postoperative pain and behavioural recovery<br>Secondary outcome measures – Negative affect and Length of stay in hospital | **Number of included studies:** 115<br>**Key characteristics:** 105 studies from 115 papers (representing 10,302 participants) were included in this review. The publication dates of the included studies ranged from 1970 to 2014 and studies were conducted in a wide range of countries. The primary studies originated from the USA (n = 36), **UK (n = 13)**, Canada (n = 9), China (n = 7), Australia (n = 6), The Netherlands (n = 5), Germany (n = 4), in Sweden (n = 3), and had one or two studies each from Austria, Brazil, Denmark, Egypt, France, India, Iran, Ireland, Italy, New Zealand, Nigeria, Romania, Serbia, Singapore, Spain, Switzerland, Taiwan and Turkey | High | **Postoperative pain**<br>Sixty one studies assessed the outcome postoperative pain; however it was only possible to include data for 38 studies with analysis of 2713 participants' data (26% of 10,302 participants randomized across all studies), in the meta-analysis. Overall, the pooled effect size (SMD) was -0.20 (95% confidence interval (CI) -0.35 to -0.06), suggesting a statistically significant effect in favour of the intervention groups. There were, however, high levels of statistical heterogeneity between studies ($I^2$ statistic = 71%).<br>**Behavioural recovery**<br>Fourteen studies (13% of 105 studies) were included that measured a behavioural recovery outcome, in which 1135 participants were randomized (11% of 10,302 participants randomized across all studies). It was not possible to combine numerical findings for behavioural recovery because few studies provided sufficient details and studies used different ways of measuring how quickly people returned to usual activities. Narratively reviewed evidence for the outcome behavioural recovery provided very low quality evidence that psychological preparation, in particular behavioural instruction, may have potential to improve behavioural recovery outcomes, but no clear conclusions could be reached.<br>**Negative affect**<br>Fifty studies reported the outcome negative affect, however only 31 studies (30% of 105 studies) could be<br>included in the meta-analysis with data from 2496 participants analysed (24% of 10,302 participants randomized across all studies). Overall, there was evidence of lower negative affect in the intervention groups compared with the control groups (SMD -0.35, 95% CI -0.54 to -0.16). Although there were very high levels of statistical heterogeneity ($I^2$ statistic = 81%).<br>**Length of stay in hospital**<br>Of the 58 studies with length of stay as an outcome, sufficient data were available for meta-analysis in 36 (34% of 105 studies: with data from 3313 participants (32% of 10,302 participants randomized across all studies). Overall, when considering all types of psychological intervention, there was evidence of shorter length of stay in the intervention groups compared with the control groups (mean difference (MD) -0.52 days, 95% CI -0.82 to -0.22) (Analysis 1.2; Figure 6). There were, however, high levels of statistical heterogeneity between studies ($I^2$ statistic = 74%).<br>**Notes**<br>Funnel plots showed no clear evidence of publication bias. |
| Tong et al. (2020) Effect of preoperative psychological interventions on elective orthopaedic surgery outcomes: a systematic review and meta-analysis. ANZ Journal of | **Intervention Type:** Psychological interventions<br>**Review period:** 1960 to January 2018<br>**Review purpose:** To investigate the types and effectiveness of preoperative psychological interventions in elective orthopaedic surgery<br>**Included study designs:** Prospective controlled clinical trials | **Number of included studies:** 19<br>**Key characteristics:**<br>Of the 19 included studies, 15 were RCTs and four were non-RCTs. Combined, they comprised 1893 patients, with mean age of 54.0 years (range 10–90), 1162 (61.4%) females and 671 (35.4%) males (no information on sex of 60 patients). Countries of included studies were not described | Moderate | **Pain**<br>Sixteen studies reported on the outcome of pain. In the acute postoperative period, five studies found a statistically significant reduction in pain scores in intervention groups and nine found no significant effect. Data from seven studies were pooled for a meta-analysis examining the outcome of pain in the acute postoperative period. Analysis did not find enough evidence to confirm reduction in postoperative pain (seven studies, 666 patients; $g = -0.15$ (95% CI -0.42, 0.13), $P = 0.305$). There was significant moderate heterogeneity ($I^2 = 59.42$, $P = 0.02$) and no evidence of publication bias based on Egger's regression analysis ($P = 0.34$). |
| Surgery, 90(3), pp.230-236. | **Included outcome measures:** Pain, anxiety, quality of life and disability and function. |  |  | **Anxiety**<br>Fourteen studies reported on postoperative anxiety, with three studies finding statistically significant reductions acutely in the intervention group, and trials unable to find a statistically significant effect between groups.<br>Data from six studies (589 patients) were pooled for meta-analysis. A moderate, statistically significant effect was demonstrated in favour of the psychological intervention ($g = -0.26$ ($-0.49, -0.03$), $P = 0.024$), with non-significant low heterogeneity ($I^2 = 29.46$, $P = 0.21$). There was no evidence of publication bias based on Egger's regression analysis ($P = 0.60$).<br>**Quality of life**<br>Two studies, one utilizing guided imagery and the other emotional counselling, measured quality of life using the Short Form Health Questionnaire (SF-36), assessing physical and mental components of this outcome at both subacute and longer-term time points. Pooled data from these studies (282 patients) showed a significant improvement in the mental component of quality of life at longer term follow-up ($g = 0.25$ ($0.02, 0.49$), $P = 0.034$), with very low heterogeneity ($I^2 = 0.00$, $P = 0.460$).<br>There was no statistically significant difference at the subacute time point ($g = 0.52$ ($-0.29, 1.33$), $P = 0.209$) with significant heterogeneity ($I^2 = 89.72$, $P = 0.002$). Similarly, no significant effect was detected for physical components of quality of life at subacute and chronic time points ($g = -0.16$ ($-0.39, 0.08$), $P = 0.189$ and $g = 0.08$ ($-0.15, 0.32$), $P = 0.485$, respectively), with non-significant low heterogeneity ($I^2 = 0.00$, $P = 0.46$ and $0.88$, respectively).<br>**Function and disability**<br>Five studies reported on measures of function and disability; all found statistically significant benefits with<br>intervention. Unfortunately, heterogeneity in measurement and reporting precluded meta-analysis. In the acute period, two studies reported statistically significant improvements in mobility and function. Another trial reported statistically significant improvements in similar functional measures for both subacute and longer-term time points. Two studies reported significant improvements in disability outcomes at subacute and longer-term follow-up, respectively.<br>**Notes**<br>Risk of publication bias assessed for pain and anxiety. |
| Multicomponent interventions |  |  |  |  |
| Citation | Review details | Included studies | Quality | Findings and observations/notes |
| Moyer et al (2017) The value of preoperative exercise and education for patients undergoing total hip and knee arthroplasty: a systematic review and meta-analysis. JBJS reviews, 5(12), p.e2. | **Intervention Type:** Preoperative exercise and education<br>**Review period:** Inception of databases to May 2016<br>**Review purpose:** To determine the longitudinal effects and efficacy of prehabilitation on postoperative outcomes in patients undergoing total hip arthroplasty (THA) or total knee arthroplasty (TKA).<br>**Included study designs:** RCTs<br>**Included outcome measures:**<br>Primary outcome measures - Patient-reported measure of pain and function.<br>Secondary outcome measures - postoperative measures of quadriceps and hamstring strength, anxiety, and length of stay. | **Number of included studies:** 35<br>**Key characteristics:**<br>Thirty five RCTs representing 2,956 participants were included in the review. A total of 1,151 underwent TKA (mean age [and standard deviation], 67 +/- 3.2 years; 419 male, 732 female), 1,193 underwent THA (69 +/- 6.4 years; 535 male, 658 female), and the joint (hip or knee) was not specified for 615 patients (68 +/- 2.8 years; 190 male, 425 female) in five RCTs. Preoperative programmes typically ran between 4 and 8 weeks (minimum, 2 weeks; maximum, 24 weeks) and 1 to 5 times per week (mean, approximately 3 times per week). Countries of included studies were not described | Low | **Pain**<br>Twenty five studies reported the effects of prehabilitation on postoperative pain. Overall, there was a small, significant difference favouring prehabilitation for improving pain (SMD = 0.14, p = 0.007). The funnel plot was symmetric and negative for the existence of publication bias (intercept = -0.17, p = 0.779). When analyzed by joint, there was a similar but non-significant SMD in patients treated for knee osteoarthritis (SMD = 0.11, p = 0.136), and a larger SMD in patients with hip osteoarthritis (SMD = 0.15, p = 0.017); the latter represented a significant improvement in pain in patients who underwent prehabilitation compared with controls.<br>**Function**<br>Twenty three studies reported the effects of prehabilitation on postoperative function. Overall, there was a moderate, significant difference favoring prehabilitation for improving function (SMD = 0.34, p < 0.001). The funnel plot was symmetric but positive for significant publication bias (intercept = 2.49, p = 0.011).<br>**Quadriceps strength** |
|  |  |  |  | Eight studies reported the effects of prehabilitation on postoperative quadriceps strength. Overall, there was a moderate, significant difference favoring prehabilitation for improving quadriceps strength (SMD = 0.29, $p = 0.042$). The funnel plot was symmetric and negative for publication bias (intercept = 0.51, $p = 0.813$). Heterogeneity was significant ($I^2 = 48.55\%$, $p = 0.049$); this was reduced when patients with hip and knee osteoarthritis were evaluated separately. The SMD was larger and significant in patients with knee osteoarthritis (SMD = 0.42, $p = 0.002$), but not significant in patients with hip osteoarthritis (SMD = 20.28, $p = 0.241$). There were too few studies reporting quadriceps strength for further THA subgroup analyses.<br>**Hamstring strength**<br>Three studies reported hamstring strength; these included only patients with knee osteoarthritis. Overall, there was a large but non-significant difference favoring prehabilitation for improving hamstring strength (SMD = 1.32, $p = 0.132$). The funnel plot was symmetric and negative for publication bias (intercept = 12.27, $p = 0.165$). Heterogeneity was significant ($I^2 = 95.91\%$, $p < 0.001$). After removing a single outlier<sup>66</sup>, the SMD decreased from 1.32 to 0.14, and heterogeneity was no longer significant ($I^2 = 11.63\%$, $p = 0.287$). There were too few studies reporting hamstring strength for further subgroup analyses.<br>**Anxiety**<br>Six studies reported the effects of prehabilitation on postoperative anxiety. Overall, there was a small, non-significant difference favoring prehabilitation for improving postoperative anxiety (SMD = 0.06, $p = 0.723$). The funnel plot was symmetric and negative for publication bias (intercept = 1.97, 95% CI = -7.54 to 11.48, $p = 0.596$). The SMDs were similar in magnitude but in opposite directions for patients with knee osteoarthritis (SMD = -0.23, $p = 0.492$) and hip<br>osteoarthritis (SMD = 0.17, $p = 0.128$). Heterogeneity was large and significant ($I^2 = 59.99\%$, $p = 0.029$), and likely due to the heterogeneity among TKA studies ($I^2 = 76.95\%$, $p = 0.037$). There were too few studies reporting anxiety for planned TKA subgroup analyses.<br>When analyzed with respect to intervention effectiveness, the SMD for effective programs in THA patients was small and non-significant (SMD = 0.20, $p = 0.178$). Only 1 THA study had an ineffective program, which was therefore not analyzed further. Both studies involving TKA reported ineffective programs for patients with knee osteoarthritis. The SMD for anxiety in these studies did not favor prehabilitation (SMD = -0.23, $p = 0.492$) and had high heterogeneity ($I^2 = 76.95\%$, $p = 0.037$), but planned subgroup analyses could not be completed because the number of studies was too small.<br>**Length of stay**<br>Nineteen studies reported the effects of prehabilitation on postoperative length of stay. Overall, there was a small-to-moderate, significant difference favoring prehabilitation (SMD = 0.37, $p = 0.001$) for decreasing hospital length of stay. The funnel plot was symmetric and negative for publication bias (intercept = 0.58, 95% CI, -2.62 to 3.77, $p = 0.707$). Heterogeneity was significant ($p < 0.001$). When THA and TKA were evaluated separately, there was a similar SMD in patients with hip osteoarthritis (SMD = 0.31, $p = 0.027$) and a higher SMD in patients with knee osteoarthritis (SMD = 0.54, $p < 0.001$); heterogeneity remained elevated ($p < 0.001$). Neither effective nor ineffective programs significantly improved the hospital length of stay for patients with hip osteoarthritis ($p = 0.237$ and $0.057$). However, for patients with knee osteoarthritis, the decrease in length of stay was moderate and significant for both effective (SMD = 0.57, $p = 0.012$) and ineffective programs (SMD = 0.52, $p = 0.036$). |
|  |  |  |  | **Notes**<br>Funnel plots for pain, quadriceps strength and hamstring strength were symmetric and negative for publication bias. |
| Yau et al. (2021) <u>Effect of preparative rehabilitation on recovery after cardiac surgery: A systematic review - ClinicalKey</u> *Annals of Physical and Rehabilitation Medicine*, 64(2), p.101391. | **Intervention Type:** Physical prehabilitation exercise and education<br>**Review period:** Up to 20 June 2019<br>**Review purpose:** To investigate the effectiveness of physical prehabilitation programs before cardiac surgery on postoperative recovery and other perioperative outcomes.<br>**Included study designs:** RCTs<br>**Included outcome measures:**<br>Primary outcomes - Perioperative mortality, postoperative complications and quality of recovery<br>Secondary outcomes included -<br>Physical or functional performance (e.g., exercise capacity, functional capacity, and physical activity level), psychosocial status (e.g., anxiety and depression levels, and quality of life), frailty level and healthcare utilisation (e.g., intensive care unit [ICU] and hospital LOS, healthcare expenditure, and cardiac rehabilitation enrolment). | **Number of included studies:** 7<br>**Key characteristics:**<br>Seven RCTs (Representing 726 participants) were included in this review. Trials were conducted in Australia, Brazil, Canada, Taiwan and the United Kingdom. The median number of participants per study was 60 (IQR 26-204) and the percentage of females was 20%. Most studies involved patients undergoing isolated coronary artery bypass graft surgery. The seven primary studies originated from Canada (n = 3), and had **one study each in each from the UK, Australia, Brazil and Taiwan** | Low | **Perioperative mortality**<br>Four studies reported the postoperative incidence of death. Whether prehabilitation reduced the risk of mortality was uncertain (Peto odds ratio [OR] 1.30, 95% confidence interval [CI] 0.28-5.95; I<sup>2</sup> = 0%; 4 studies, 532 participants; low-certainty evidence)<br>**Postoperative complications**<br>Findings were uncertain as to whether prehabilitation reduced the risk of stroke (RR 0.59, 95% CI 0.08-4.56; I<sup>2</sup> = 0%; 2 studies, 230 participants; low-certainty evidence) and atrial fibrillation (RR 0.75, 95% CI 0.38 to 1.46; I<sup>2</sup> = 50%; 4 studies, 214 participants; very low-certainty evidence).<br>Low-certainty evidence was found for the risk of acute kidney injury (RR 2.60, 95% CI 0.12 to 58.48; 1 study, 26 participants), conduction disturbance (RR 0.29, 95% CI 0.01 to 6.50) and heart valve infection (RR 3.12, 95% CI 0.13 to 75.67), so the effect of prehabilitation was unclear.<br>Findings were uncertain as to whether prehabilitation reduced the risk of postoperative pneumonia (RR 0.38, 95% CI 0.01 to 14.64; I<sup>2</sup> = 67%; 2 studies, 82 participants) and atelectasis (RR 0.74, 95% CI 0.04 to 14.21; I<sup>2</sup> = 71%, 2 studies, 71 participants); all very low-certainty evidence. Prehabilitation probably reduced the risk of pleural effusion (RR 0.43, 95% CI 0.19 to 0.97; 1 study, 56 participants; moderate-certainty evidence).<br>There was unclear evidence of the effect of prehabilitation on risk of a gastrointestinal event (RR |
|  |  |  |  | 0.86, 95% CI 0.06 to 12.28; 1 study, 26 participants) and reoperation (RR 0.29, 95% CI 0.01 to 6.50) because of low-certainty evidence. Low-certainty evidence was found for the association between prehabilitation and risk reduction of delayed extubation (> 24 hr.), and its effect was unclear (RR 0.17, 95% CI 0.01 to 3.29; 1 study, 26 participants). Another study reported that prehabilitation might slightly reduce the time to endotracheal extubation for patients receiving phase 1 cardiac rehabilitation before surgery (MD -286 min, 95% CI -572.1 to 0.1; 1 study, 56 participants; low certainty evidence), but its effect was inconclusive.<br>**Quality of recovery**<br>None of the included trials measured this outcome with validated and reliable questionnaires. Therefore, the effect of prehabilitation on quality of recovery was unclear.<br>**Physical or functional performance**<br>**- Functional capacity**<br>The mean change in functional capacity with the 6-min walk test (6MWT) was assessed between baseline and the immediate preoperative period in 2 trials. Whether prehabilitation improved total distance walked during the 6MWT measured immediately before surgery was uncertain (MD 81.9, 95% CI -23.3 to 187.1; 2 studies, 41 participants; very low-certainty evidence), with high heterogeneity ($I^2 = 92\%$) possibly because of differences in duration of prehabilitation programs. Data were not pooled because when 6MWT was measured in the postoperative period substantially differed in the remaining studies<br>**- Physical activity level**<br>Findings were uncertain as to whether prehabilitation improved MVPA10-min (MD 2 min/week, 95% CI -29.3 to 33.3; 1 study, 23 participants) or Total PAspor (MD 125 min/week, 95% CI -35.5 to 285.5) between baseline and the immediate preoperative period [43], and Total |
|  |  |  |  | PAspor (MD -91 min/week, 95% CI -700 to 518; 1 study, 13 participants) between baseline and 3 months postoperatively [42], all low-certainty evidence.<br>**Psychosocial status**<br>**- Anxiety level**<br>Four trials assessed the change in anxiety level at various times with different outcome measures. Data provided by 3 trials were insufficient for analysis.<br>**- Depression**<br>Meta analysis was not performed<br>**- Quality of life**<br>Findings were unclear as to whether physical prehabilitation improved the Medical Outcomes Study Short-Form 36 (SF-36) physical component summary (MD 1.37, 95% CI -1.87 to 4.61; I<sup>2</sup> = 80%; 2 studies, 337 participants) and mental component summary (MD 1.06, 95% CI -3.73 to 5.86; I<sup>2</sup> = 86%; 2 studies, 337 participants) immediately before surgery; all provided low certainty evidence [39,41]. We found low-certainty evidence with an unclear effect of prehabilitation on the SF-36 sub-scores of the physical component summary (MD -0.20, 95% CI -2.31 to 1.91; 1 study, 117 participants) and mental component summary (MD 1.20, 95% CI -1.47 to 3.87) between baseline and 6 weeks postoperatively<br>**Frailty level**<br>Frailty was assessed in one trial with the Functional Frailty Index (FFI) and modified Fried Phenotype (m-FP). Findings were uncertain as to the effect of prehabilitation on mean changes in FFI score (MD 0.02, 95% CI -0.02 to 0.06; 26 participants) and m-FP (MD -0.10 deficits, 95% CI -0.77 to 0.57) from baseline to the immediate preoperative period, all low-certainty evidence.<br>**Healthcare utilisation**<br>**- Length of stay** |
|  |  |  |  | Overall, the effect of prehabilitation and ICU LOS was unclear (MD 1.41 hr., 95% CI -7.97 to 10.78; I<sup>2</sup> = 74%; 5 studies, 377 participants; low-certainty evidence). Patients participating in PPPs had slightly shorter postoperative hospital LOS (MD -0.62 days, 95% CI -0.93 to -0.32; I<sup>2</sup> = 0%; 2 studies, 235 participants; moderate-certainty evidence). Physical prehabilitation may slightly reduce total hospital LOS (MD -0.66 days, 95% CI -1.29 to -0.03; I<sup>2</sup> = 45%; 4 studies, 567 participants; low-certainty evidence).<br>**- Healthcare expenditure**<br>Two trials assessed the preoperative healthcare expenditure. One trial estimated that the net cost savings associated with prehabilitation was approximately \$133 CAD/ patient/day. Another trial evaluated healthcare expenditure (including costs of general practitioner visits and hospital admission) and quality-adjusted life years (QALYs) in the 8 weeks before surgery. The authors found no difference in mean cost between prehabilitation and control groups (MD £1.73, 95% CI - 17.73 to 20.60; 204 participants). Patients with prehabilitation had similar QALYs as controls (MD 0.006 QALYs, 95% CI -0.002 to 0.015; 204 participants). In the cost-acceptability curve, prehabilitation was likely (>90% probability) cost-effective in terms of £/QALY.<br>**- Postoperative cardiac rehabilitation enrolment**<br>Findings were uncertain as to whether prehabilitation improved postoperative cardiac rehabilitation enrolment (RR 1.42, 95% CI 0.87-2.30, I<sup>2</sup> = 44%; 2 studies, 235 participant; very low certainty evidence).<br>**Notes**<br>A sensitivity analysis of low risk of bias trials to estimate the robustness of results was planned, but all trials were rated as high risk because of blinding issues. We were unable to perform subgroup analyses on the effect of prehabilitation duration or assess publication bias<br>because of fewer than 10 trials meeting the inclusion criteria. |
| Education-based interventions |  |  |  |  |
| Citation (Country) | Review details | Included studies | Quality | Findings and observations/notes |
| Burgess et al. (2019) <u>The effect of preoperative education on psychological, clinical and economic outcomes in elective spinal surgery: a systematic review</u>. In Healthcare (Vol. 7, No. 1, p. 48). Multidisciplinary Digital Publishing Institute. | **Intervention Type:** Preoperative education<br>**Review period:** Up to July 2018<br>**Review purpose:** To determine whether a preoperative education intervention improves psychological, clinical or economic outcomes from elective spinal surgery<br>**Included study designs:** RCTs<br>**Included outcome measures:** Clinical (Self-reported and performance based), Psychological and Economic evaluations | **Number of included studies:** 7<br>**Key characteristics:**<br>The search yielded seven RCTs (11 papers) (664 participants) that investigated the effect of preoperative education interventions on psychological, clinical and economic outcomes following elective spine surgery. Countries of included studies were not described | Low | There is limited, fair-quality evidence that supports the inclusion of a preoperative education session for improving clinical (pain, function and disability), economic (quality-adjusted life years, healthcare expenditure, direct and indirect costs) and psychological outcomes (anxiety, depression and fear-avoidance beliefs) from spinal surgery. Other benefits are reported to be improved patient knowledge, feelings of better preparation, reduced negative thinking and increased levels of physical activity after the intervention. No differences in quality of life, return to work, physical indicators or postoperative complications were reported. From the limited evidence, it is not possible to conclusively recommend that preoperative education should be delivered as a standalone intervention before elective spine surgery; however, given the low risk profile and promising benefits, future research in this area is warranted.<br>**Notes**<br>One study was scored as “high quality” and the remaining as “fair quality”. Consistently low scoring items included the blinding of participants and personnel, blinding of outcome assessment and other bias. However, due to the nature of the intervention delivered, it was not always possible to blind the patient and clinician, and therefore no study was considered “poor quality”, which would suggest an important methodological limitation that could invalidate results. |
| Ng et al. (2021) <u>The</u> | **Intervention Type:** Education | **Number of included studies:** 22 | Low | **Anxiety**<br>**Preoperative anxiety** |
| effectiveness of preoperative education interventions on improving perioperative outcomes of adult patients undergoing cardiac surgery: a systematic review and meta-analysis.<br>Eur J Cardiovasc Nurs. 2021 Dec 29;zvab123. doi: 10.1093/eurjcn/zvab123. Epub ahead of print. PMID: 34964470. | **Review period:** Inception of databases until Dec 2020<br>**Review purpose:** To synthesize the best available evidence evaluating the effectiveness of preoperative education in improving perioperative outcomes of patients undergoing cardiac surgery.<br>**Included study designs:** RCTs<br>**Included outcome measures:**<br>**Primary outcome measures** - participants' anxiety, depression and knowledge.<br>**Secondary outcomes** - participants' pain intensity, pain interference with daily activities, postoperative complications, length of hospitalisation, length of ICU stay, satisfaction with the intervention and care, and HRQoL. | **Key characteristics:**<br>This review included 22 RCTs conducted from 1987 till 2020 and involved 3167 participants (2503 males, 664 females). Sample sizes ranged from 20 to 406, with participants' mean age between 52.0 and 72.1-years. All studies had mixed-gender groups except three trials being males-only. The primary studies originated from Canada (n = 5), China (n = 3), United Kingdom (n = 3), United States (n = 3), Iran (n = 2), and had one study each from Austria, Brazil, Germany, Hong Kong, Norway, and Turkey. |  | Six studies involving 657 participants examining post intervention preoperative anxiety were pooled in a meta-analysis. Preoperative education yielded statistically significant results, with a large effect size for post intervention preoperative anxiety reduction [SMD = -0.94, 95% CI (-1.74 to -0.14), Z= 2.31, P= 0.02], as compared to usual care. Significant heterogeneity was detected (I<sup>2</sup> = 95%, X<sup>2</sup> = 102.76, P< 0.00001).<br>**Postoperative anxiety**<br>Six studies involving 623 participants examining post intervention postoperative anxiety with adequate data were pooled.<br>The overall interventional effect on postoperative anxiety reduction was statistically significant, with a small to moderate effect size [SMD = -0.49, 95% CI (-0.71 to -0.26), Z = 4.25, P< 0.0001], compared to usual care. Notably, moderate heterogeneity was reported (I<sup>2</sup> = 43%, x<sup>2</sup> = 8.80, P= 0.12).<br>**Depression**<br>Meta-analysis was conducted for three studies assessing depression and involved 397 participants. Preoperative education displayed significantly lowered depression scores of a small effect size [SMD = -0.21, 95% CI (-0.41 to -0.02), Z = 2.13, P = 0.03], favouring intervention than usual care.<br>**Knowledge**<br>Two studies involving 110 participants were pooled for the meta-analysis of knowledge. Those in intervention group had statistically significant improved knowledge, with a large effect size [SMD = 2.59, 95% CI (2.07–3.10), Z = 9.81, P < 0.00001]. Heterogeneity that might not be important was indicated (I<sup>2</sup> = 0%, x<sup>2</sup> = 0.47, P = 0.49).<br>**Secondary outcomes** |
|  |  |  |  | Meta-analyses examined the effectiveness of preoperative education on seven remaining secondary outcomes compared to usual care. The intervention group participants had significantly improved satisfaction with the education intervention and care [SMD = 0.46, 95% CI (0.02–0.89), Z = 2.05, P= 0.04, I<sup>2</sup> = 30%] and reduced length of ICU stay [MD = -3.26, 95% CI (-6.02 to -0.50), Z = 2.32, P= 0.02]. While effect size was large for the length of ICU stay, moderate heterogeneity was observed (I<sup>2</sup> = 44%, P= 0.10). Additionally, meta-analyses revealed no statistically significant group differences observed for the outcomes of: pain intensity, pain interference with daily activities, postoperative complications, length of hospitalisation, and HRQoL (P ≥ 0.05)<br>**Notes**<br>A funnel plot was produced for the length of hospitalisation, involving 12 studies. Egger's test suggested no publication bias was detected (Z = 0.0433, P = 0.9654) |
| Van der Gucht et al. (2021) <u>Effectiveness of preoperative pain science education on pain, psychological factors and physical functioning: A systematic review</u>. Clinical Rehabilitation, 35(10), pp.1364-1382. | **Intervention Type:** Education (pain science)<br>**Review period:** Up to February 28, 2021<br>**Review purpose:** To synthesize the evidence on the effectiveness of pain science education on pain, psychological factors and physical functioning in adults who underwent surgery.<br>**Included study designs:** Randomised clinical trials and controlled clinical trials | **Number of included studies:** 9 publications reporting 8 studies<br>**Key characteristics:** Seven RCTs and one controlled clinical trial were included. Sample sizes ranged from 31 to 402 with a total of 1,078 unique participants, 468 of whom received pain science education. The primary studies originated from the USA (n = 5), Denmark (n = 2), Spain (n = 1) and Chile (n = 1) | Moderate | Only two RCTs showed significant interaction effects.<br>**Psychological factors**<br>Psychological factors (pain catastrophizing and kinesiophobia) decreased in participants who had received pain science education before total knee arthroplasty, while this was not the case in the control group (P-value < 0.001, $\eta^2$p:0.11). The RCT showing a significant interaction effect for postoperative pain was focused on breast cancer patients and therefore not within the scope of our review.<br>**Physical functioning**<br>No statistically significant interaction effects were reported for outcomes on physical functioning<br>**Notes** |
|  | **Included outcome measures:**<br>Pain, Psychological factors and/or<br>Physical functioning |  |  | Vast majority of included studies (n=6) were at high risk of bias. The only study reporting on postoperative pain outcomes was focused on breast cancer patients, therefore findings are not eligible for use in this rapid review. |

\* Overall rating for the review based on an assessment using the AMSTAR 2 critical appraisal tool (Shea et al. 2017)

Of the final 17 systematic reviews, 14 combined their results in a meta-analysis while three used a narrative synthesis method to report their data. Search dates ranged from 2014 to 2021, and primary studies included in the systematic reviews were mostly randomised controlled trials. The most common elective surgery type studied was orthopaedic surgery (n = 9). Others included cardiac surgery (n = 3), and cardiac and abdominal surgery (n = 2). Three reviews did not have a specific surgical focus. The majority of included systematic reviews focused on exercise-based interventions (n = 9). Other interventions were educational (n = 3), psychological (n = 2), and smoking cessation interventions (n = 1). Two reviews focused on multicomponent interventions. Narrative summaries of the evidence are presented below based on types of interventions.

The methodological quality of the 17 prioritised systematic reviews were assessed using the AMSTAR assessment tool. Most were rated low quality (n = 11), one was rated critically low, two rated as moderate and three rated as high quality (Table 2). The moderate and low quality systematic reviews generally failed to adequately investigate the possible impact of risk of bias on the findings. Most authors commented that included primary studies had small sample sizes and studies often varied by surgical type. The trials also often had blinding issues.

### 2.2 Exercise-based interventions

Nine systematic reviews (Alshewaier et al., 2017, Blasco et al., 2021, Fenton et al., 2021, Husted et al., 2020, Katsura et al., 2015, Potts et al., 2022, Rodrigues et al., 2021, Wang et al., 2021, Wang et al., 2016) provided evidence on the effectiveness of preoperative exercise interventions on improving outcomes of patients awaiting elective surgery. Six of the systematic reviews focussed on orthopaedic surgical procedures (three total knee replacement/arthroplasty, two anterior cruciate ligament reconstruction, and one on both total knee replacement and total hip replacement); two systematic reviews on cardiac and abdominal surgery, and one on cardiac surgery. With regards to interventions, three of the nine systematic reviews examining the evidence of exercised-based interventions, were focussed specifically on preoperative rehabilitation (prehabilitation) (Alshewaier et al., 2017, Fenton et al., 2021, Wang et al., 2016); two systematic reviews focused on preoperative inspiratory muscle training (IMT) (Katsura et al., 2015, Rodrigues et al., 2021), and one systematic review focused on sensorimotor training (Blasco et al., 2021). The remaining three systematic reviews focussed on preoperative strength based exercises (Husted et al., 2020, Potts et al., 2022, Wang et al., 2021). Outcome measures were either preoperative or early postoperative outcomes and included those specific to orthopaedic surgery: pain, muscle strength, functional activity, performance-based function, balance, range of motion (ROM), and those specific to cardiac and abdominal surgery: postoperative respiratory parameters, postoperative pulmonary complications (PPCs), and change in aneurysm size. Other non-surgery specific outcomes of interest included quality of life, length of hospital/intensive care unit stay, 30-day mortality, readmission, cost-analysis, and adverse events. The considerable variation (heterogeneity) and methodological limitations across included studies may compromise the applicability of these findings.

Fenton et al. (2021) found that prehabilitation exercise **may reduce cardiac and renal complications in people awaiting abdominal aortic aneurysm (AAA) repair, compared with usual care (no exercise)**. However, it is uncertain whether prehabilitation exercise reduces the occurrence of 30-day mortality, pulmonary complications, need for re-intervention, or postoperative bleeding post-AAA repair.

Wang et al. (2016) found that prehabilitation may **slightly improve early postoperative pain (4 weeks or less), function, and time to resume activities of daily living (climbing stairs, toilet use, chair use)** in patients planning to undergo joint replacement surgery. Effects were found to be similar for both knee and hip replacement surgery, but did not affect outcomes such as length of stay, quality of life and costs.

Alshewaier et al. (2017) found that preoperative physiotherapy rehabilitation **is effective for improving treatment outcomes following anterior cruciate ligament injury, including increasing knee-related function and improving muscle strength**. No significant differences in pain, quality of life, and range of motion were found between control and intervention groups. Physical function outcomes were mixed across included studies.

Rodrigues et al. (2021) found that preoperative breathing therapy through IMT on patients undergoing cardiac surgery may help **improve respiratory performance after surgery, reduce postoperative pulmonary complications and length of hospital stay**.

Katsura et al. (2015) found that preoperative IMT was **associated with a reduction of postoperative atelectasis, pneumonia, and duration of hospital stay in adults undergoing cardiac and major abdominal surgery**. No significant differences in quality of life, maximal inspiratory muscle strength, mechanical ventilation exceeding 48 hours, and all-cause mortality within 30 days, were found between study groups.

Blasco et al. (2021) found **limited evidence suggesting that preoperative sensorimotor training enhanced preoperative self-reported function, functional performance, knee function and pain compared with conventional care**. Benefits were only maintained in terms of functional performance up to three months after surgery. Functional performance was found to be similar after one year, regardless of whether such training is implemented.

Potts et al. (2021) found that **four to 16 weeks of preoperative exercise could significantly increase quadriceps strength preoperatively in patients with unilateral anterior cruciate ligament injury**, but any persistent postoperative strength benefit from undertaking a standardised preoperative intervention is unclear.

Wang et al. (2021) found that a preoperative exercise intervention before total knee arthroplasty (TKA) **can significantly improve knee ROM flexion and flexibility, reduce inflammatory pain and stiffness, improve muscle strength, improve joint function**, and thus improve the quality of life of patients.

Husted et al. (2020) found **no relationship between preoperative knee extensor exercise dosage and change in knee-extensor strength, knee pain, or physical function prior to and following TKA**. Preoperative exercise including knee-extensor muscle strength exercise increased knee-extensor strength moderately prior to, but not three months following TKA.

#### 2.2.1 Bottom line results for exercise interventions

The evidence suggests that the implementation of preoperative exercise interventions, including prehabilitation, could help improve preoperative and postoperative outcomes such as pain, muscle strength and function, and reduced incidence of postoperative complications, in people awaiting elective surgery. However, considerable variation (heterogeneity) and methodological limitations across included studies may compromise the applicability of these findings.

### 2.3 Educational interventions

Three systematic reviews (Burgess et al., 2019, Ng et al., 2021, Van der Gucht et al., 2021) provided evidence on the effectiveness of preoperative educational interventions on improving outcomes of patients awaiting elective surgery. One of the reviews targeted patients awaiting orthopaedic surgery (spinal surgery), another on cardiac surgery, and the third review did not have any specific surgical focus (elective surgery). Outcomes sought included participants’ pre- and postoperative anxiety, depression, knowledge, pain, physical functioning, postoperative complications, length of hospitalisation, quality of life, and cost analysis.

Burgess et al. (2019) found limited, fair quality evidence supporting the inclusion of preoperative education sessions for improving clinical (preoperative and postoperative pain, function and disability), economic (quality-adjusted life years, healthcare expenditure, direct and indirect costs) and psychological outcomes (anxiety, depression and fear-avoidance beliefs) from spinal surgery. The review authors believe that from the limited evidence, it is not possible to conclusively recommend the delivery of preoperative education as a standalone intervention before elective spine surgery.

Ng et al. (2021) found that **preoperative education had large significant effects on reducing post intervention preoperative anxiety, length of ICU stay, and improving knowledge** in adult patients undergoing cardiac surgery. Small significant effects were also found for reducing postoperative anxiety, depression, and enhancing satisfaction.

Van der Gucht et al. (2021) found that **preoperative pain science education did not result in significant effects on pain, psychological factors or physical functioning compared to controls**, in patients waiting to undergo total knee arthroplasty.

#### 2.3.1 Bottom line results for educational interventions

Evidence suggests that preoperative educational interventions are effective at improving knowledge in patients awaiting elective surgery. However, the evidence supporting the use of these interventions in improving pre- and postoperative pain and physical functioning in orthopaedic patients is limited. In addition, the evidence provided mixed results for the effectiveness of preoperative educational interventions on psychological outcomes.

### 2.4 Psychological interventions

Two systematic reviews (Powell et al., 2016, Tong et al., 2020) provided evidence on the effectiveness of preoperative psychological interventions on improving outcomes for patients awaiting elective surgical procedures. One review targeted elective orthopaedic patients undergoing a psychological intervention initiated prior to elective orthopaedic surgery (Tong et al., 2020), while the other targeted adult participants undergoing elective surgery under general anaesthesia (Powell et al., 2016). Outcome measures sought included pain, anxiety, quality of life, length of hospital stay, physical function, and negative affect.

Tong et al. (2020) found that psychological interventions had a **moderate statistically significant effect on postoperative anxiety and improved the mental component of quality of life at longer term follow-up**. Meta-analysis did not find enough evidence to confirm a reduction in postoperative pain.

Powell et al. (2016) found that psychological interventions might be beneficial for the outcomes postoperative pain, behavioural recovery, negative affect and length of stay, and is unlikely to be harmful. However, caution must be exercised when interpreting the results because of heterogeneity in the types of surgery, interventions and outcomes.

#### 2.4.1 Bottom line results for psychological interventions

The evidence surrounding preoperative psychological interventions is limited, but indicates it may have a positive effect on anxiety and mental health components of quality of life postoperatively. However, the evidence in support of such interventions in reducing postoperative pain is inconclusive.

### 2.5 Smoking cessation

One systematic review (Thomsen et al., 2014) provided evidence on the effectiveness of preoperative smoking interventions (intensive and brief) on smoking cessation in patients awaiting elective surgery. This review found that **preoperative smoking interventions providing behavioural support and offering nicotine replacement therapy (NRT) increased short-term smoking cessation and may reduce postoperative morbidity**. Intensive preoperative smoking cessation interventions appear to reduce the incidence of postoperative complications, but not brief interventions. Similar effects were found for wound complication, but no studies detected significant differences in duration of hospital admission or postoperative pulmonary or cardiovascular complications.

### 2.6 Multicomponent interventions

Two systematic reviews (Moyer et al., 2017, Yau et al., 2021) provided evidence on the effectiveness of preoperative multicomponent interventions on improving outcomes of patients awaiting elective surgery. Both systematic reviews investigated the effectiveness of interventions consisting of a form of exercise combined with education. One of the reviews targeted orthopaedic patients undergoing total hip arthroplasty (THA) or total knee arthroplasty (TKA), while the other targeted adults undergoing cardiac surgery. Outcomes sought included patient-reported measures of pain and function, psychosocial status, hospital length of stay, postoperative complications, and perioperative mortality.

Yau et al. (2021) found uncertain effects of a multicomponent intervention consisting of physical prehabilitation and education, on postoperative clinical outcomes, including perioperative mortality and postoperative atrial fibrillation, and physical activity levels. However, these interventions may improve postoperative functional capacity and slightly shorten hospital length of stay after cardiac surgery. Authors also identified no difference in mean cost between prehabilitation and control groups, and patients with prehabilitation had similar QALYs as controlled. In the cost-acceptability curve, prehabilitation was likely (>90% probability) cost-effective in terms of £/QALY.

Moyer et al. (2017) found that **in patients undergoing TKA, significant improvements were observed in function, quadriceps strength, and length of stay**, after the delivery of a preoperative programme involving exercise and education. **In patients undergoing THA, significant improvements were observed in pain, function, and length of stay**. No significant difference was identified between the intervention and control group in terms of anxiety.

#### 2.6.1 Bottom line results for multicomponent interventions

Evidence suggests that preoperative interventions consisting of both exercise and education could shorten the length of hospital stay as well as improvements in postoperative pain, function, and muscle strength.

## 3. DISCUSSION

### 3.1 Summary of the findings

There is evidence to suggest that preoperative interventions including exercise, education, smoking cessation, and psychological interventions are effective at improving selected preoperative and postoperative outcomes in people awaiting elective surgery. Delivering these interventions together as part of a multicomponent intervention may also be beneficial for postoperative outcomes. However, considerable variation (heterogeneity) and methodological limitations across included primary studies, as well as differences in the types of elective surgeries reviewed, may compromise the applicability of these findings.

### 3.2 Limitations of the available evidence

Most of the evidence identified in this report were derived from orthopaedic surgery reviews – some of which sought outcome measures specific to orthopaedic surgery. It is unclear whether the findings from such reviews can be applied to other surgical fields.

This rapid review did not identify any evidence relating to the use of social prescribing or other community-centred approaches to support surgical wait-listed patients.

There was limited data on the socio-demographic characteristics of the patients recruited in the included reviews. Gender was reported in only eight reviews, with the majority showing a male predominance. Data on race, ethnicity, income, and education were not reported, and would prove valuable in giving an accurate understanding of the impact of these interventions on wait-listed patients.

None of the systematic reviews identified in this rapid review measured the quality of recovery (postoperative experience) or surgical treatment outcomes. In addition, the outcomes of feeling prepared for surgery, return to work, postoperative patient knowledge and postoperative behavioural recovery outcomes were each examined by a single systematic review.

In addition, a limited number of systematic reviews considered post intervention, preoperative outcomes so we cannot tell if these interventions help to turn the ‘waiting list’ into a ‘preparation list’, enabling patients to be fully supported in using the waiting period proactively.

The ten ongoing systematic reviews report an intention to examine preoperative exercise interventions, prehabilitation and smoking cessation interventions across various surgical disciplines. Although most report they will be looking at outcomes already identified in our included systematic reviews, they may also identify additional outcomes that will contribute to the existing evidence base.

No evidence was identified in the context of the current COVID-19 pandemic. It is unclear what impact the pandemic (and any associated restrictions) could have on the conduct of these interventions.

### 3.3 Implications for policy and practice

This report has highlighted the benefits of preoperative interventions for patients awaiting elective surgery. The current evidence supports the use of exercise, education, smoking cessation, and psychological interventions to support and improve the outcomes of wait-listed patients. Policymakers and clinicians should consider incorporating such interventions into health professionals’ curriculums.

The effect of social prescribing interventions in supporting patients awaiting surgery needs to be established. In addition, further research is required to understand how various patient subgroups respond to preoperative interventions, including those from underserved and minority ethnic groups.

### 3.4 Strengths and limitations of this Rapid Review

Sources included in this rapid review were identified through an extensive search conducted between the 7^th^ and 8^th^ of February 2022. Additional searches were also conducted on 17^th^ February 2022 to identify systematic reviews on social prescribing. Full text screening was peer reviewed by two independent researchers. The data extraction was performed by one researcher and two independent researchers carried out consistency checking.

The quality assessment of the included systematic reviews was undertaken using AMSTAR 2, a specific and validated tool to assess the quality of systematic reviews. Quality assessment was conducted by one reviewer and consistency checked by a second reviewer. Quality assessment findings (the overall rating for each review) are outlined in Table 2. Authors of identified records (including protocols for relevant systematic reviews) were contacted to obtain information or progress reports about their publications and other relevant references.

Although we have made efforts to capture all relevant publications and reduce the risk of bias, it is possible that additional eligible publications may have been missed or we may have introduced some biases in this review.

Due to time constraints, we did not attempt to undertake any assessment of the outcomes using GRADE. Therefore, we are unable to comment on the quality of the overall body of evidence examining innovations aimed at supporting patients on elective surgical waiting lists.

## Data Availability

All data produced in the present study are available upon reasonable request to the authors

## Abbreviations

AAA: Abdominal Aortic Aneurysm
ACL: Anterior Cruciate Ligament
AMSTAR: A MeaSurement Tool to Assess systematic Reviews
BMI: Body Mass Index
CABG: Coronary Artery Bypass Grafting
CKC: Closed Kinetic Chain
COVID-19: Coronavirus disease 2019
ENT: Ear, Nose and Throat
GI: Gastrointestinal
GRADE: Grading of Recommendations, Assessment, Development and Evaluations
HOOS: The Hip Disability and Osteoarthritis Outcome Score
HRQOL: Health Related Quality of Life
HSS: Hospital for Special Surgery
ICERs: Incremental Cost-Effectiveness Ratios
ICU: Intensive Care Unit
IMT: Inspiratory Muscle Training
KOOS: The Knee Injury and Osteoarthritis Outcome Score
LOS: Length of Hospital Stay
MOS: Medical Outcomes Study
NACRT: Neoadjuvant Chemotherapy and/or Radiotherapy
NRS: Numerical Rating Scales
NRT: Nicotine Replacement Therapy
OA: Osteoarthritis
OECD: Organisation for Economic Co-operation and Development
OKC: Open Kinetic Chain
PAD: Peripheral Arterial Disease
PET: Preoperative Exercise Therapy
PPCs: Postoperative Pulmonary Complications
PPE: Preoperative Patient Education
PROMS: Patient Reported Outcome Measures
QALY: Quality-Adjusted Life Years
QoL: Quality of Life
RES: Rapid Evidence Summary
RCT: Randomised Control Trial
ROM: Range of Movement
RR: Rapid Review
SF-36: 36-Item Short-Form Health Survey
SR: Systematic review
THA: Total Hip Arthroplasty
THR: Total Hip Replacement
TKA: Total Knee Arthroplasty
TKR: Total Knee Replacement
UK: United Kingdom
USA: United States of America
VAS: Visual Analogue Scales
WCEC: Welsh Covid-19 Evidence Centre
WHYMPI: West Haven-Yale Multidimensional Pain Inventory
WOMAC: Western Ontario McMaster Universities Osteoarthritis Index

## 5. RAPID REVIEW METHODS

### 5.1 Prior Rapid Evidence Summary

As an initial stage for this rapid review, a Rapid Evidence Summary (RES) was conducted in February 2022. The RES represents a preliminary review of the literature that is used to clarify the research question and needs of the requestor, gauge the potential size of the available literature, inform the methods and design of the subsequent rapid review, and provide limited interim findings to the stakeholder. It is based on a search of key resources and the assessment of abstracts. Priority is given to studies representing robust evidence synthesis. No quality appraisal or evidence synthesis are conducted.

The RES addressed the slightly boarder question of “What are the effectiveness of innovations/alternative treatments to support patients on surgical waiting lists?” This included interventions to support surgical patients on the two types of waiting lists i.e. waiting to see the specialist (time to decision) and those awaiting surgery (time to treatment), as well as alternative treatments to avoid or reduce the need for surgery. The findings were fed back to the stakeholders, and it was decided that the Rapid Review should focus on patients awaiting surgery (time to treatment) as the RES indicated no evidence was available for patients waiting for surgical opinion. It was also decided that due to limited bariatric surgical services in Wales, patients awaiting bariatric surgical services would be excluded from the review. Finally, outcome measures would include both preoperative and postoperative measures.

### 5.2 Eligibility criteria

The following eligibility criteria were used to identify studies for inclusion in the rapid review:

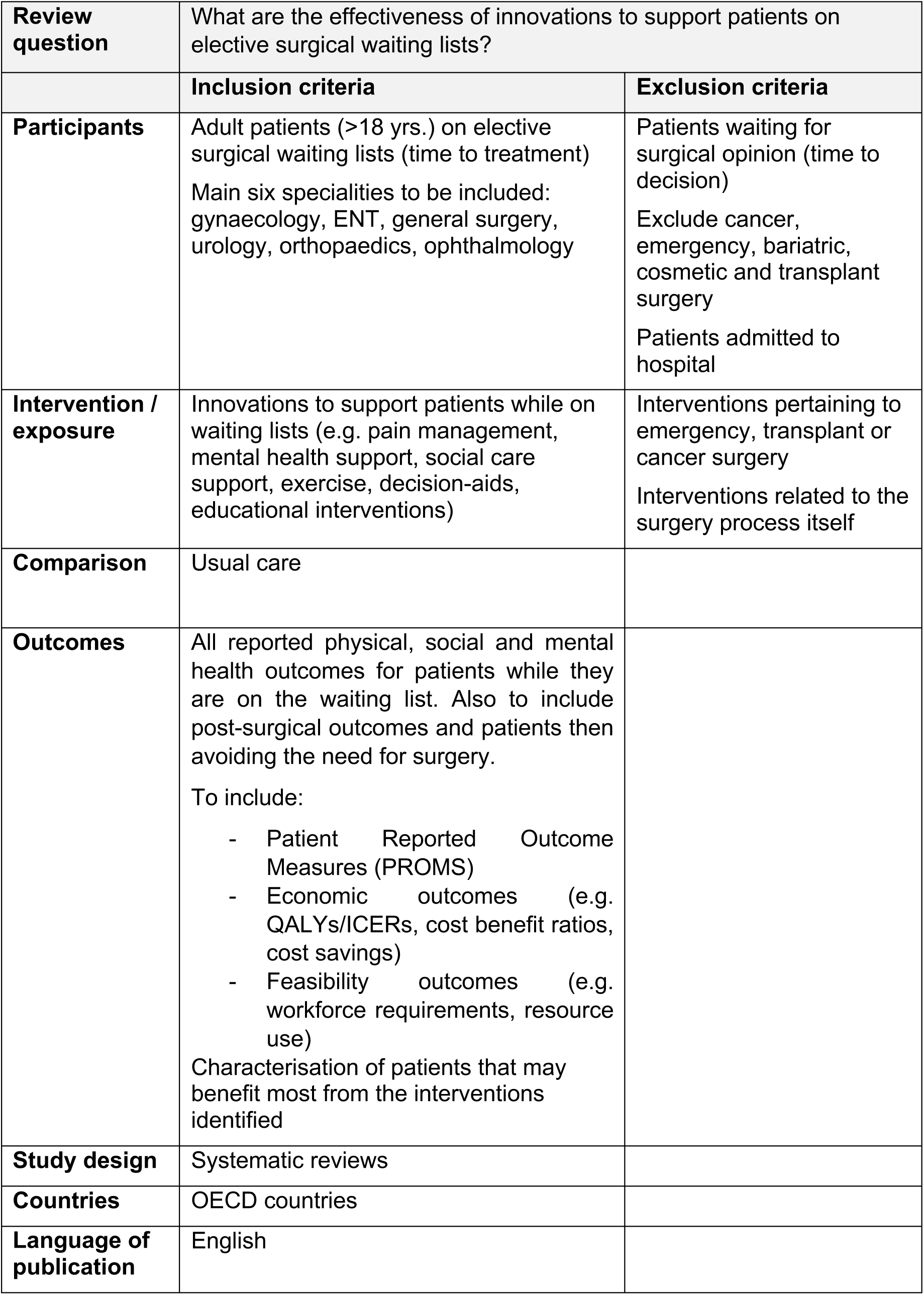
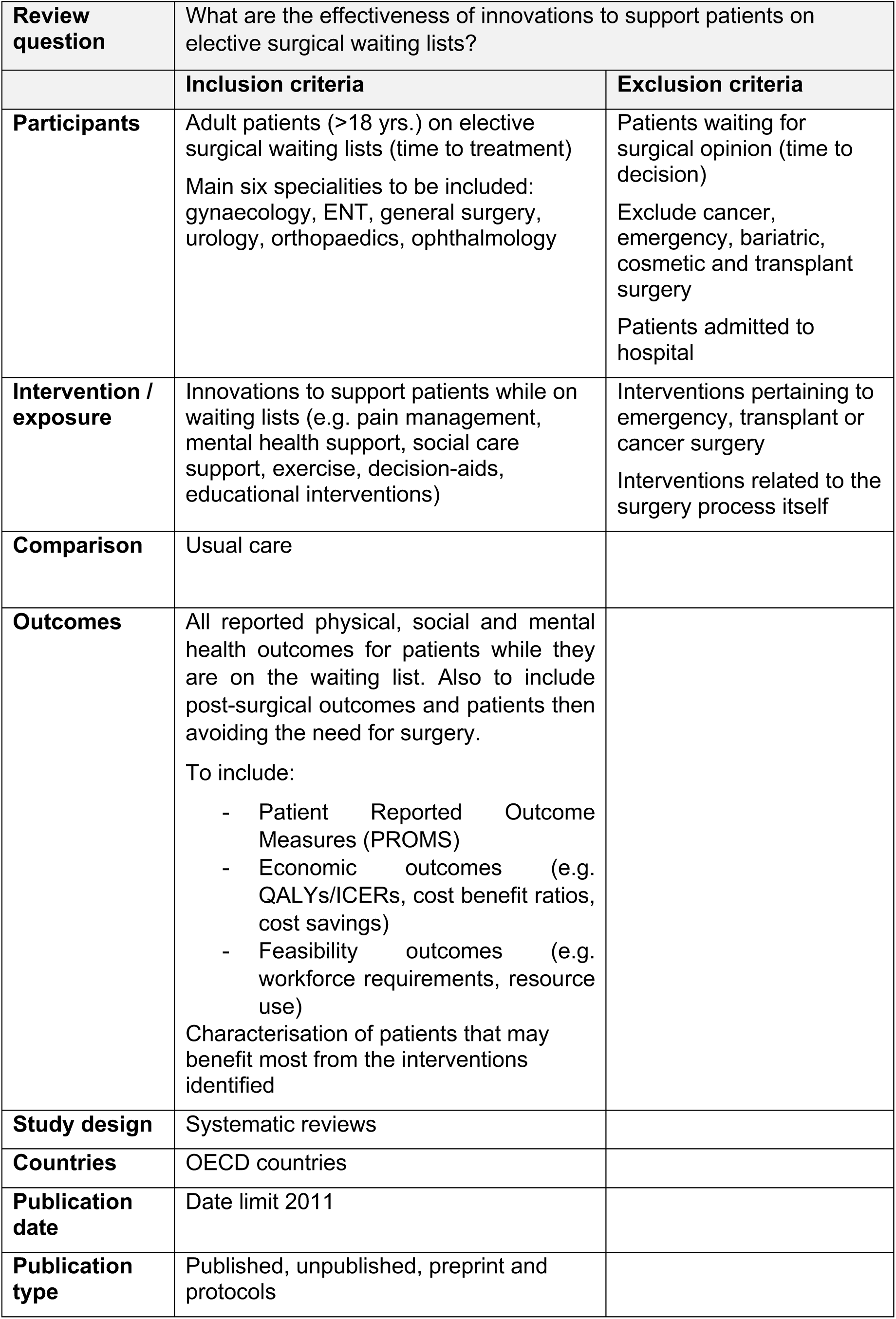

| **Review question** | What are the effectiveness of innovations to support patients on elective surgical waiting lists? |  |
| --- | --- | --- |
|  | **Inclusion criteria** | **Exclusion criteria** |
| **Participants** | Adult patients (>18 yrs.) on elective surgical waiting lists (time to treatment)<br>Main six specialities to be included: gynaecology, ENT, general surgery, urology, orthopaedics, ophthalmology | Patients waiting for surgical opinion (time to decision)<br>Exclude cancer, emergency, bariatric, cosmetic and transplant surgery<br>Patients admitted to hospital |
| **Intervention / exposure** | Innovations to support patients while on waiting lists (e.g. pain management, mental health support, social care support, exercise, decision-aids, educational interventions) | Interventions pertaining to emergency, transplant or cancer surgery<br>Interventions related to the surgery process itself |
| **Comparison** | Usual care |  |
| **Outcomes** | All reported physical, social and mental health outcomes for patients while they are on the waiting list. Also to include post-surgical outcomes and patients then avoiding the need for surgery.<br>To include:<br>- Patient Reported Outcome Measures (PROMS)<br>- Economic outcomes (e.g. QALYs/ICERs, cost benefit ratios, cost savings)<br>- Feasibility outcomes (e.g. workforce requirements, resource use)<br>Characterisation of patients that may benefit most from the interventions identified |  |
| **Study design** | Systematic reviews |  |
| **Countries** | OECD countries |  |
| **Language of publication** | English |  |
| **Review question** | What are the effectiveness of innovations to support patients on elective surgical waiting lists? |  |
|  | **Inclusion criteria** | **Exclusion criteria** |
| **Participants** | Adult patients (>18 yrs.) on elective surgical waiting lists (time to treatment)<br>Main six specialities to be included: gynaecology, ENT, general surgery, urology, orthopaedics, ophthalmology | Patients waiting for surgical opinion (time to decision)<br>Exclude cancer, emergency, bariatric, cosmetic and transplant surgery<br>Patients admitted to hospital |
| **Intervention / exposure** | Innovations to support patients while on waiting lists (e.g. pain management, mental health support, social care support, exercise, decision-aids, educational interventions) | Interventions pertaining to emergency, transplant or cancer surgery<br>Interventions related to the surgery process itself |
| **Comparison** | Usual care |  |
| **Outcomes** | All reported physical, social and mental health outcomes for patients while they are on the waiting list. Also to include post-surgical outcomes and patients then avoiding the need for surgery.<br>To include:<br>- Patient Reported Outcome Measures (PROMS)<br>- Economic outcomes (e.g. QALYs/ICERs, cost benefit ratios, cost savings)<br>- Feasibility outcomes (e.g. workforce requirements, resource use)<br>Characterisation of patients that may benefit most from the interventions identified |  |
| **Study design** | Systematic reviews |  |
| **Countries** | OECD countries |  |
| **Publication date** | Date limit 2011 |  |
| **Publication type** | Published, unpublished, preprint and protocols |  |

### 5.3 Literature search

As part of the initial Rapid Evidence Summary, COVID-19 specific and general repositories of evidence reviews were systematically searched between the 7^th^ and 8^th^ of February 2022, for relevant sources. Searches were limited to English-language publications published after 2011, and did not include searches for primary studies.

A list of resources searched can be found below:

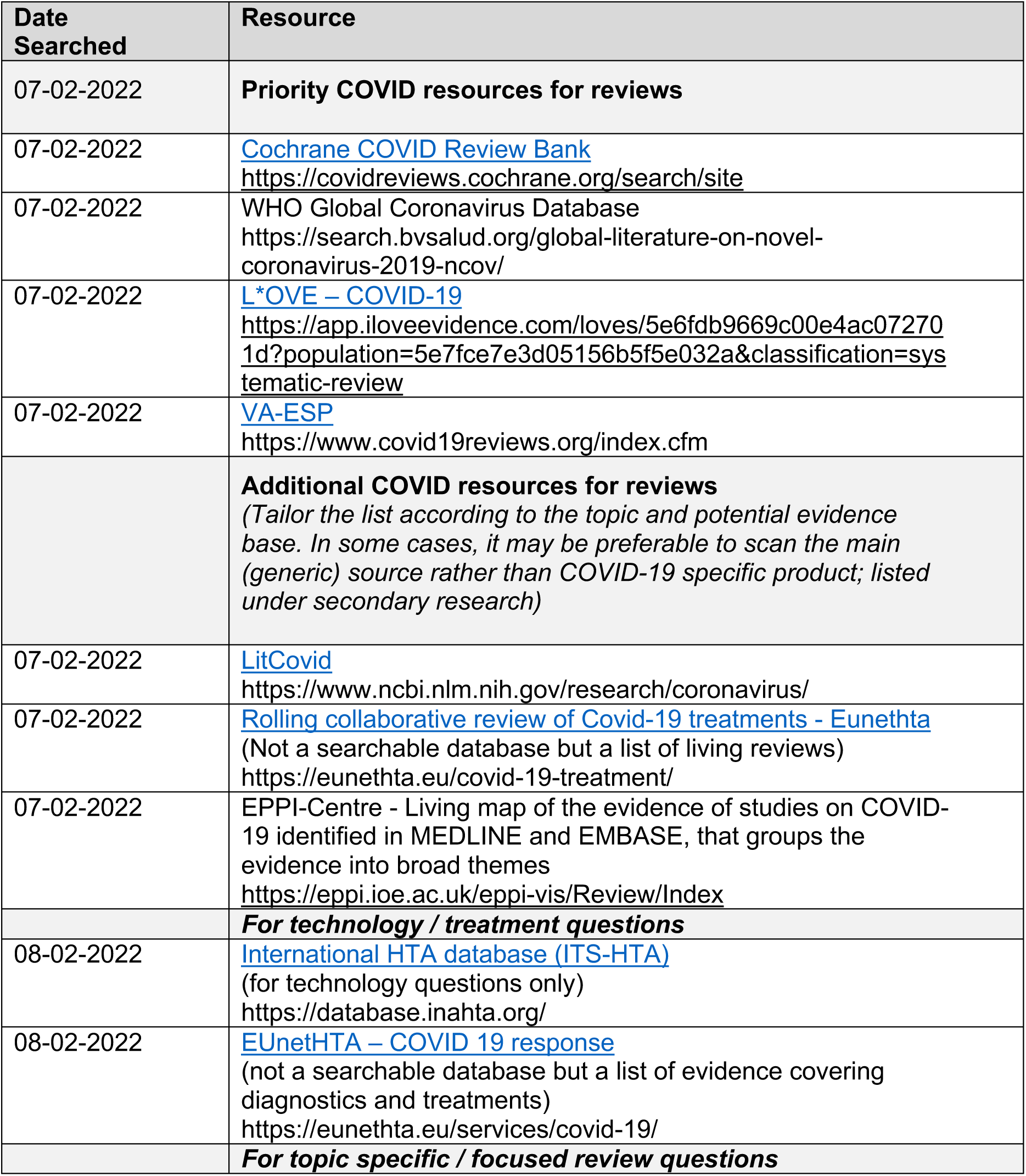
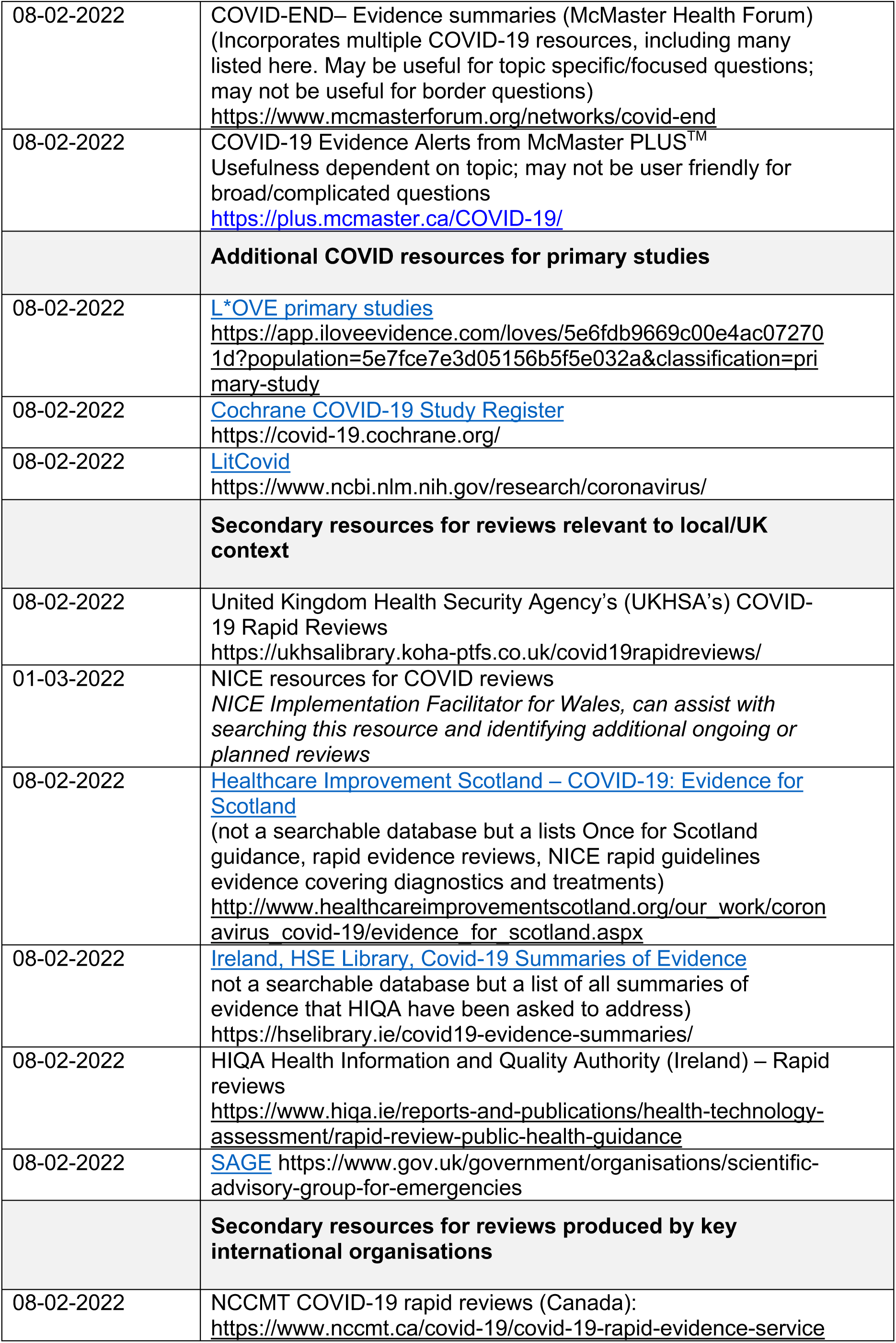
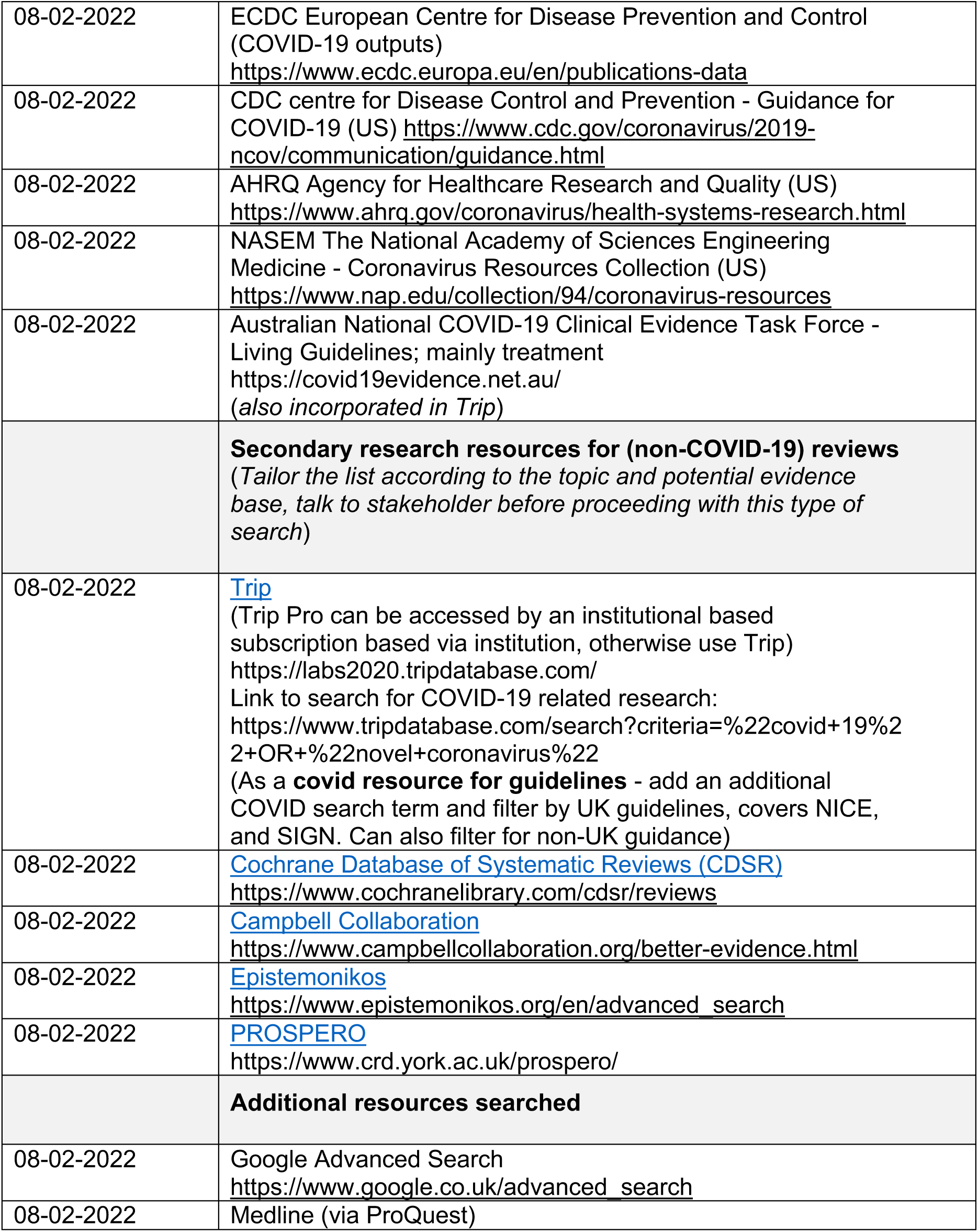

An information specialist devised and conducted the searches using the concepts: ‘Patients waiting for surgery’, ‘interventions to support patients whilst waiting (e.g. pain management, mental health support, social care support)’, ‘secondary and tertiary research’. The searches combined free text words and descriptors where available. The search strategy used for MEDLINE is available in Appendix 1. An additional search was conducted in MEDLINE to identify systematic reviews on social prescribing (Appendix 2). The authors of systematic reviews protocols identified were contacted to check the status of their publications.

### 5.4 Study selection process

The searches yielded a total of 1,098 records. Records were imported into an Endnote database and duplicates were removed. After deduplication, a total of 887 records remained. We excluded 228 records prior to screening because they were published prior to 2011, or because their titles contained the words cancer, children, emergency or transplant. A total of 659 records were screened for inclusion, independently in duplicate, by three reviewers using the title and abstract. Disagreements were resolved through discussion. A total of 140 records were then screened at full text, independently in duplicate, by three reviewers. Where there was disagreement, they were resolved through discussion. A total of 48 systematic reviews and 10 protocols met our inclusion criteria.

#### 5.4.1 Prioritisation process

Full text screening resulted in a large number of included studies, as such, a prioritisation process was used to reduce the number of studies and allow for an in-depth review. Initially, using predetermined criteria, we examined the methodology of the included systematic reviews to determine if the included sources could be considered systematic reviews. These criteria included: whether or not a systematic search had been conducted (e.g. more than one database with examples of keywords or search strategies used); whether the study selection process involved at least two reviewers and had been clearly outlined; and whether quality appraisal of included studies had been conducted. Additional characteristics including the review focus, intervention type, characteristics of included studies and recorded outcomes were also reviewed. At this stage a total of six studies were determined not to have met the criteria to be considered a systematic review and were therefore not prioritised. The remaining 42 systematic reviews were then grouped by intervention type and compared to determine which of them would be prioritised and included in the synthesis of this Rapid Review. Prioritisation was based on multiple factors, including the recency of the review, comprehensiveness, quality of the review and the number of included studies, as well as the overlap of included studies with other, more recent or relevant systematic reviews. After applying the prioritisation criteria, 17 systematic reviews were included for critical appraisal (section 5.7), data extraction and synthesis. The data extraction for the 31 studies not prioritised, and reasons for this, can be seen in Appendix 3.

### 5.5 Study selection flow chart

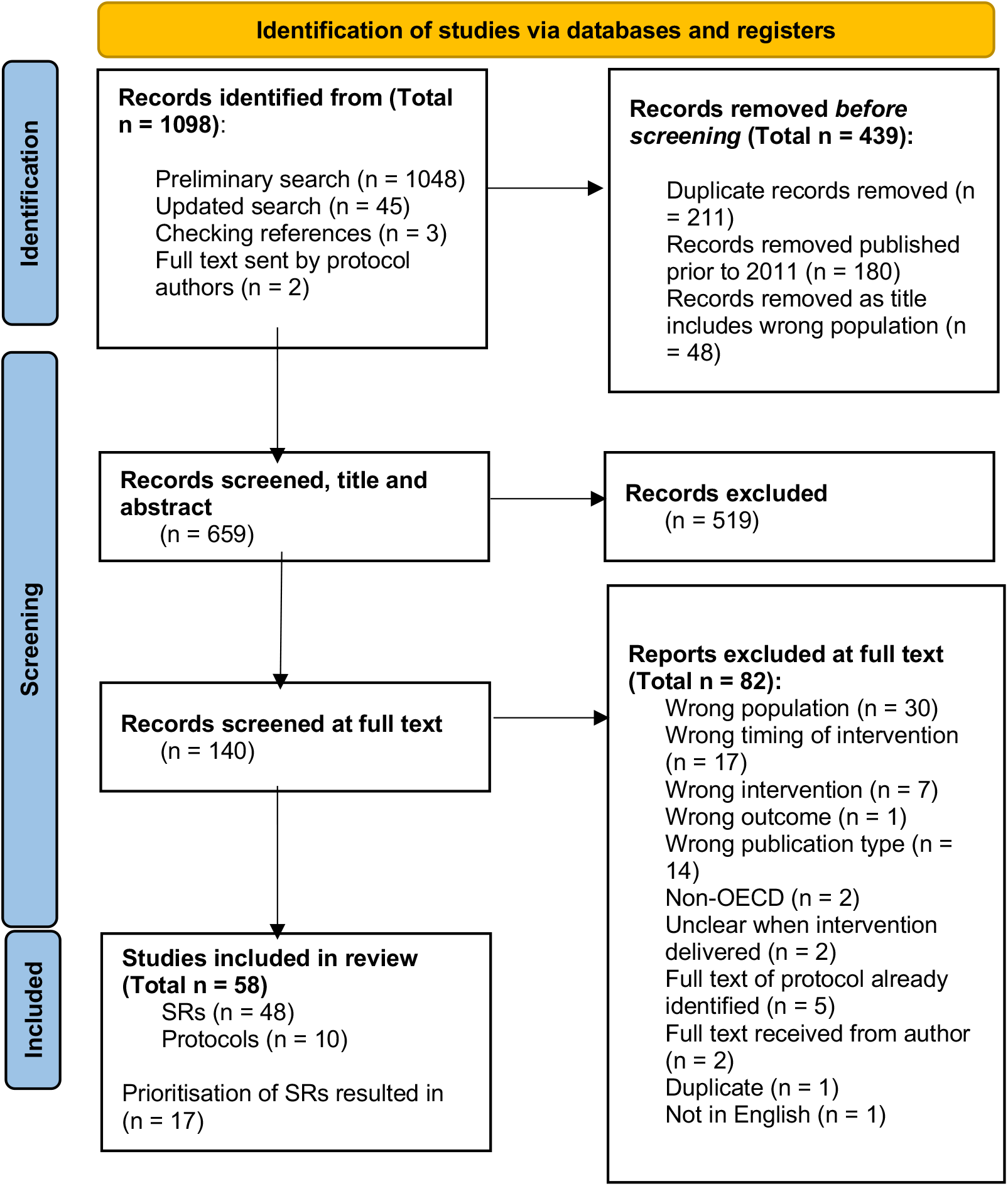
*From:* Page MJ, McKenzie JE, Bossuyt PM, Boutron I, Hoffmann TC, Mulrow CD, et al. The PRISMA 2020 statement: an updated guideline for reporting systematic reviews. BMJ 2021;372:n71. doi: 10.1136/bmj.n71

For more information, visit: http://www.prisma-statement.org/

### 5.6 Data extraction

Data were extracted by two reviewers and included demographic and outcome data. The following information was extracted from each secondary study:

- Title
- Author
- Year
- Country of included studies
- Intervention type
- Review period
- Review purpose
- Included study design
- Included outcome measures
- No of included studies
- Key characteristics
- Findings and observations

### 5.7 Quality appraisal

Quality assessment was undertaken by a single reviewer, with verification of all judgements by a second reviewer. Any discrepancies were discussed and resolved amongst the review team. Critical appraisal was conducted using AMSTAR 2, a valid tool for assessing the quality of systematic reviews of randomised controlled trials (Shea et al., 2017). The reviewers agreed to give prominence to seven critical domains (see below) that could affect the validity of a review and its conclusions.

Critical domains:

- Protocol registered before commencement of the review (item 2)
- Adequacy of the literature search (item 4)
- Justification for excluding individual studies (item 7)
- Risk of bias from individual studies being included in the review (item 9)
- Appropriateness of meta-analytical methods (item 11)
- Consideration of risk of bias when interpreting the results of the review (item 13)
- Assessment of presence and likely impact of publication bias (item 15)

Consistency checking for study selection and data extraction was not considered critical as these elements had already been considered as part of the prioritisation process. The results (overall rating) from the quality appraisal can be seen in Table 2.

### 5.8 Synthesis

A map outlining interventions and outcomes assessed by the studies included within all 48 systematic reviews meeting our inclusion criteria was created in order to visualise the breadth of the evidence identified and the evidence gaps (Table 1). A narrative synthesis was conducted reporting results from the 17 prioritised systematic reviews.

## 6.2 Acknowledgements

The authors would like to thank Phil Coles, Olivia Shorrocks, Caroline Mills, Brendan Collins, Luke Davies, Mark Davies, Ruth Crowder, Tracey Breheny and Nathan Davies for their contributions during stakeholder meetings in guiding the focus of the review and interpretation of findings.

## 7. ABOUT THE WALES COVID-19 EVIDENCE CENTRE (WCEC)

The WCEC integrates with worldwide efforts to synthesise and mobilise knowledge from research.

We operate with a core team as part of Health and Care Research Wales, are hosted in the Wales Centre for Primary and Emergency Care Research (PRIME), and are led by Professor Adrian Edwards of Cardiff University.

The core team of the centre works closely with collaborating partners in Health Technology Wales, Wales Centre for Evidence-Based Care, Specialist Unit for Review Evidence centre, SAIL Databank, Bangor Institute for Health & Medical Research/ Health and Care Economics Cymru, and the Public Health Wales Observatory.

Together we aim to provide around 50 reviews per year, answering the priority questions for policy and practice in Wales as we meet the demands of the pandemic and its impacts.

### Director

Professor Adrian Edwards

### Website

https://healthandcareresearchwales.org/about-research-community/wales-covid-19-evidence-centre

### All reports can be downloaded from the WCEC Library

https://healthandcareresearchwales.org/wales-covid-19-evidence-centre-report-library

## 8 APPENDIX

**Appendix 1.**
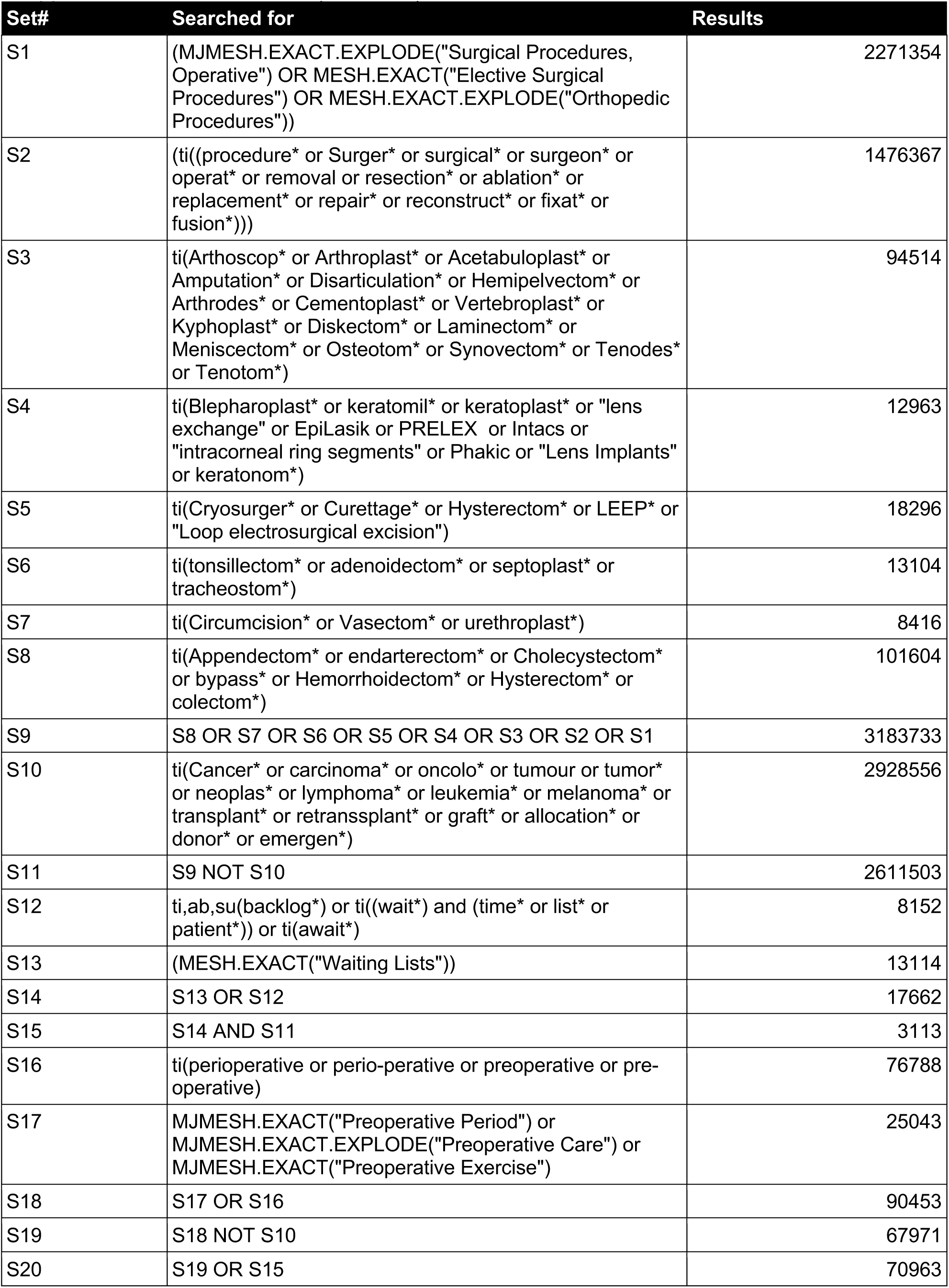
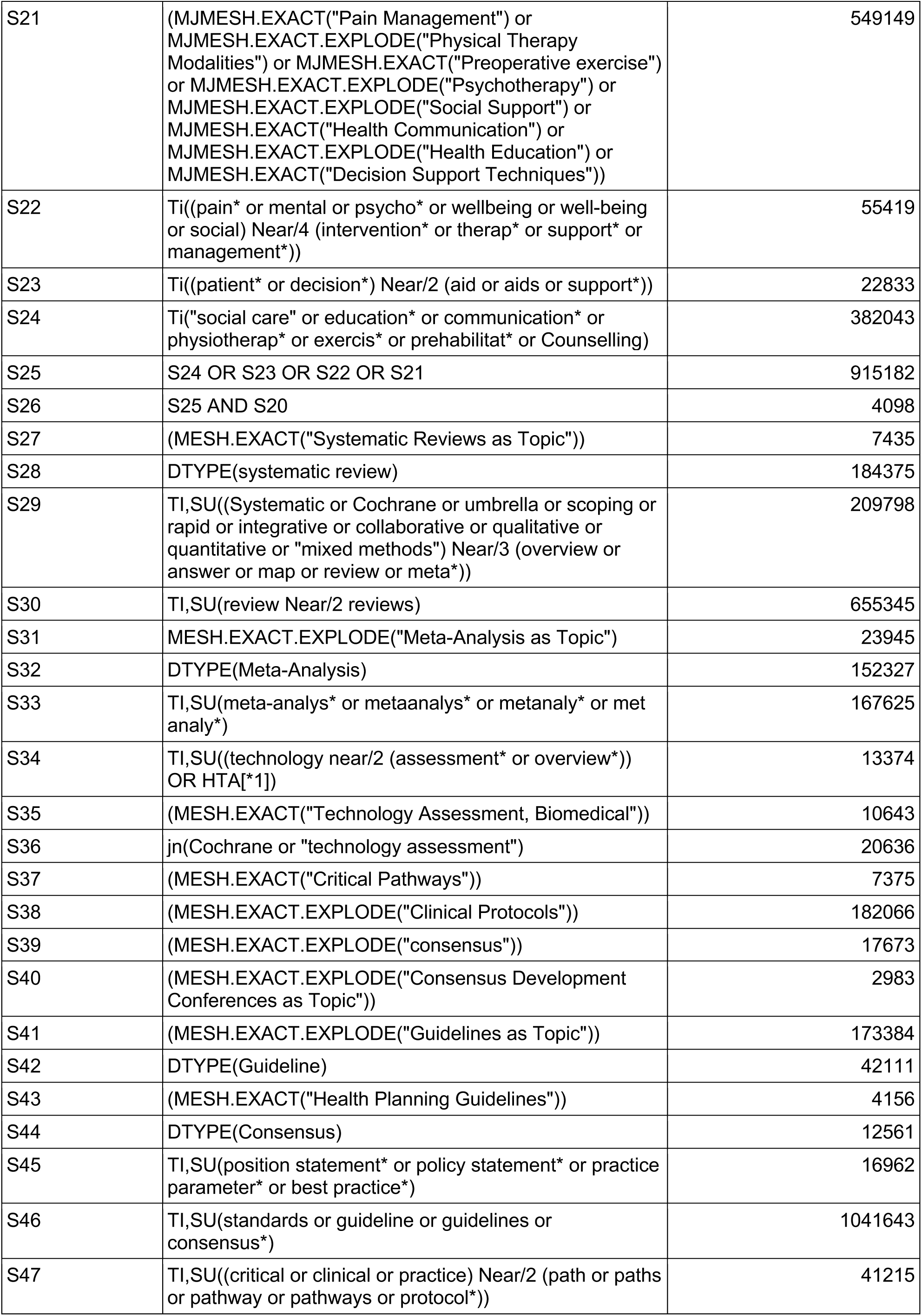
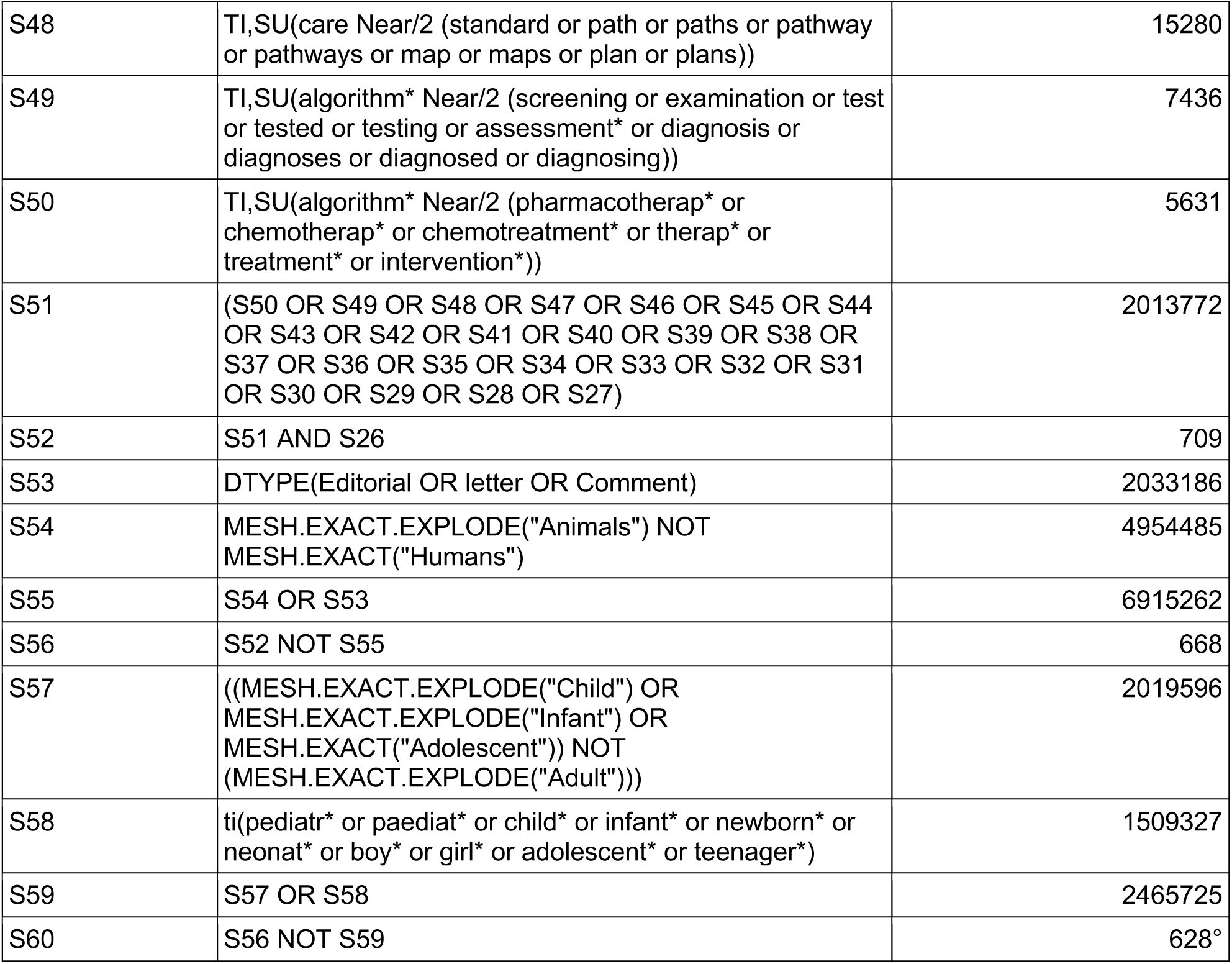
MEDLINE search (ProQuest)

**Appendix 2.** MEDLINE search (ProQuest) - Social prescribing

| Set# | Searched for | Results |
| --- | --- | --- |
| S1 | ti(social near/4 prescrib*) | 178° |
| S2 | (MESH.EXACT("Systematic Reviews as Topic")) | 7516 |
| S3 | DTYPE(systematic review) | 185184 |
| S4 | TI,SU((Systematic or Cochrane or umbrella or scoping or rapid or integrative or collaborative or qualitative or quantitative or "mixed methods") Near/3 (overview or answer or map or review or meta*)) | 211224 |
| S5 | TI,SU(review Near/2 reviews) | 657842 |
| S6 | MESH.EXACT.EXPLODE("Meta-Analysis as Topic") | 24045 |
| S7 | DTYPE(Meta-Analysis) | 152908 |
| S8 | TI,SU(meta-analys* or metaanalys* or metanaly* or met analy*) | 168448 |
| S9 | TI,SU((technology near/2 (assessment* or overview*)) OR HTA[*1]) | 13396 |
| S10 | (MESH.EXACT("Technology Assessment, Biomedical")) | 10650 |
| S11 | jn(Cochrane or "technology assessment") | 20650 |
| S12 | (MESH.EXACT("Critical Pathways")) | 7386 |
| S13 | (MESH.EXACT.EXPLODE("Clinical Protocols")) | 182366 |
| S14 | (MESH.EXACT.EXPLODE("consensus")) | 17715 |
| S15 | (MESH.EXACT.EXPLODE("Consensus Development Conferences as Topic")) | 2984 |
| S16 | (MESH.EXACT.EXPLODE("Guidelines as Topic")) | 173477 |
| S17 | DTYPE(Guideline) | 42147 |
| S18 | (MESH.EXACT("Health Planning Guidelines")) | 4157 |
| S19 | DTYPE(Consensus) | 12564 |
| S20 | TI,SU(position statement* or policy statement* or practice parameter* or best practice*) | 17003 |
| S21 | TI,SU(standards or guideline or guidelines or consensus*) | 1042575 |
| S22 | TI,SU((critical or clinical or practice) Near/2 (path or paths or pathway or pathways or protocol*)) | 41252 |
| S23 | TI,SU(care Near/2 (standard or path or paths or pathway or pathways or map or maps or plan or plans)) | 15323 |
| S24 | TI,SU(algorithm* Near/2 (screening or examination or test or tested or testing or assessment* or diagnosis or diagnoses or diagnosed or diagnosing)) | 7446 |
| S25 | TI,SU(algorithm* Near/2 (pharmacotherap* or chemotherap* or chemotreatment* or therap* or treatment* or intervention*)) | 5643 |
| S26 | MESH.EXACT("Economics") | 466321 |
| S27 | MESH.EXACT.EXPLODE("Costs and Cost Analysis") | 255186 |
| S28 | MESH.EXACT("Economics, Nursing") | 4012 |
| S29 | MESH.EXACT("Economics, Medical") | 9186 |
| S30 | MESH.EXACT("Economics, Pharmaceutical") | 3055 |
| S31 | MESH.EXACT.EXPLODE("Economics, Hospital") | 25493 |
| S32 | MESH.EXACT("Economics, Dental") | 1920 |
| S33 | MESH.EXACT.EXPLODE("Fees and Charges") | 33569 |
| S34 | MESH.EXACT.EXPLODE("Budgets") | 13969 |
| S35 | TI,SU(Budget* OR economic* or cost or costs or costly or costing or price or prices or pricing or pharmacoeconomic* or pharmaco-economic* or expenditure or expenditures or expense or expenses or financial or finance or finances or financed) | 664856 |
| S36 | TI,SU(value Near/2 (money or monetary)) | 460 |
| S37 | MESH.EXACT.EXPLODE("models, economic") | 16042 |
| S38 | (TI,SU(economic model*)) | 16224 |
| S39 | MESH.EXACT.EXPLODE("markov chains") | 15583 |
| S40 | TI,SU(markov) | 18993 |
| S41 | MESH.EXACT("monte carlo method") | 30853 |
| S42 | (TI,SU(monte carlo)) | 38650 |
| S43 | MESH.EXACT("Decision Theory") | 957 |
| S44 | (TI,SU((decision* Near/2 (tree* or analy* or model*)))) | 20209 |
| S45 | S44 OR S43 OR S42 OR S41 OR S40 OR S39 OR S38 OR S37 OR S36 OR S35 OR S34 OR S33 OR S32 OR S31 OR S30 OR S29 OR S28 OR S27 OR S26 OR S25 OR S24 OR S23 OR S22 OR S21 OR S20 OR S19 OR S18 OR S17 OR S16 OR S15 OR S14 OR S13 OR S12 OR S11 OR S10 OR S9 OR S8 OR S7 OR S6 OR S5 OR S4 OR S3 OR S2 | 2646027 |
| S46 | DTYPE(Editorial OR letter OR Comment) | 2036313 |
| S47 | MESH.EXACT.EXPLODE("Animals") NOT MESH.EXACT("Humans") | 4959437 |
| S48 | S47 OR S46 | 6923240 |
| S49 | S45 NOT S48 | 2350238* |
| S50 | S49 AND S1 | 45° |

**Appendix 3.** Included systematic reviews not prioritised

| Citation | Review details | Included studies | Comments /notes |
| --- | --- | --- | --- |
| **Multicomponent interventions** |  |  |  |
| Baimas-George et al. (2020)<br>Prehabilitation in Frail Surgical Patients: A Systematic Review<br>*World Journal of Surgery*, 44(11), pp.3668-3678. | **Intervention Type:** Multicomponent (Prehabilitation exercise, nutrition and psychosocial counseling)<br>**Review period:** up to September 2019<br>**Review purpose:** To investigate the impact of prehabilitation on postoperative outcomes in frail, surgical patients<br>**Included study designs:** RCTs, case-control, or observational studies<br>**Included outcome measures:**<br>**Primary outcome measures** - postoperative length of stay, complications and survival<br>**Secondary outcomes** – feasibility, cost-effectiveness and components of prehabilitation programmes | **Number of included studies:** 5<br>**Key characteristics:** Two RCT pilot studies, two case–control studies, and one case series. The studies were published between 2010 and 2017 and include, in total, 265 patients. Of these, 117 had colorectal disease, 51 had hip osteoarthritis, 75 had an upper gastrointestinal (GI) malignancy, and 22 had coronary or valve disease. Patient age ranged from 44 to 97 years old<br>Gender distribution of included participants not stated | Not prioritised based on included study designs |
| Milder et al. (2018)<br>The role of prehabilitation in frail surgical patients: A systematic review<br>*Acta Anaesthesiologica*<br>Wiley Online Library<br>62(10), pp.1356-1366. | **Intervention Type:** Multicomponent (Prehabilitation exercise alone, exercise and nutrition, inspiratory muscle training)<br>**Review period:** up to April 2018<br>**Review purpose:** This systematic review appraises the evidence available for prehabilitation in frail surgical patients. We proposed that exercise prehabilitation would especially benefit frail patients, with improvements in preoperative functional capacity, and reductions in complications and length of hospital stay | **Number of included studies:** 8<br>**Key characteristics:** Two of the eight included studies were ongoing. Of the six completed studies, there were two RCTs, one prospective case series, one pre-post design, one non-randomised trial and one case series. The six completed studies represented 268 participants. Age ranges of included participants not stated but all studies focused on frail patients and the term elderly was used in the search strategy. Gender distribution of included participants not stated | Not prioritised based on included study designs and overlap of studies included in other systematic reviews |
|  | **Included study designs:** RCTs, non-randomised control trials, prospective case series, case series and Pre-post designs<br>**Included outcome measures:** Feasibility, preoperative function, functional recovery, mortality, postoperative complications and length of hospital stay |  |  |
| Silkman Baker and McKeon. (2012)<br>Does Preoperative Rehabilitation Improve Patient-Based Outcomes in Persons Who Have Undergone Total Knee Arthroplasty? A Systematic Review<br>*PM&R*, 4(10), pp.756-767. | **Intervention Type:** Multicomponent (Prehabilitation - exercise, education, home adaptation)<br>**Review period:** 1950 to February 2011<br>**Review purpose:** To focus on how total knee arthroplasty (TKA) preoperative rehabilitation affects quality of life, pain, and physical outcomes after surgery<br>**Included study designs:** RCTs<br>**Included outcome measures:** Pain and function, hospital length of stay | **Number of included studies:** 7<br>**Key characteristics:** Seven randomised control trials representing 299 participants. All subjects were awaiting a primary total knee arthroplasty (TKA) as a result of OA of the knee. Mean ages of the groups were similar (control group, 68.7 years; rehabilitation group, 68.5 years), gender was similar (control group, 59.5% female; rehabilitation group, 63% female) | Not prioritised based on the year searches were conducted (2011) |
| Simmons and Smith. (2013)<br>Effectiveness of pre-operative physiotherapy-based programmes on outcomes following total knee arthroplasty: a systematic review and meta-analysis<br>*Physical Therapy Reviews*, 18(1), pp.1-10. | **Intervention Type:** Multicomponent (prehabilitation - exercise, education)<br>**Review period:** January 1990 to November 2011<br>**Review purpose:** To determine the effectiveness of preoperative physiotherapy-based interventions on outcomes following Total knee arthroplasty (TKA)<br>**Included study designs:** Randomised and non-randomised controlled trials<br>**Included outcome measures:** The primary outcome was function following TKA. Objective measures of function also included sit to stand ability, walking<br>speed/ability and stair climbing ability. Secondary outcomes assessed included: knee ROM, length of hospital stay (LOS), pain and muscle strength | **Number of included studies:** 11<br>**Key characteristics:** The total number of participants reviewed was 656 (237 males and 419 females). Mean age was 67.1 years ranging from 63 years to 72 years. All studies recruited participants with moderate to severe osteoarthritis. Two studies included five and 19 participants respectively with rheumatoid arthritis | Not prioritised based on the year searches were conducted (2011) |
| Wallis and Taylor. (2011)<br>Pre-operative interventions (non-surgical and non-pharmacological) for patients with hip or knee osteoarthritis awaiting joint replacement surgery – a systematic review and meta-analysis<br>*Osteoarthritis and cartilage*, 19(12), pp.1381-1395. | **Intervention Type:** Multicomponent (education, exercise, pain management, physiotherapy)<br>**Review period:** up to August 2010<br>**Review purpose:** To determine the effect of preoperative non-pharmacological and non-surgical interventions before and after joint replacement for patients with knee or hip osteoarthritis awaiting lower limb joint replacement surgery on pain, musculoskeletal impairment, activity, quality of life, and health service utilisation<br>**Included study designs:** RCTs<br>**Included outcome measures:** Pain, musculoskeletal impairment, activity, quality of life, and health service utilisation. Data also collected on adverse events, recruitment and adherence to the interventions | **Number of included studies:** 23<br>**Key characteristics:** From 23 RCTs involving 1,461 participants, 922 were awaiting knee replacement, 305 awaiting hip replacement and 234 awaiting either hip or knee replacement. The mean age of participants was 67.2 years and 66% were women. From available data the average body mass index was 30.2 kg/m<sup>2</sup>. The mean number of days on the waiting list was 81 days and mean duration of osteoarthritis symptoms was 6.7 years | Not prioritised based on the year searches were conducted (2010) |
| Exercise-based interventions |  |  |  |
| Citation | Review details | Included studies | Comments /notes |
| Cabilan et al. (2015)<br>The effectiveness of prehabilitation or preoperative exercise for surgical patients: a systematic review<br>*JBIC Evidence Synthesis*, 13(1), pp.146-187. | **Intervention Type:** Exercise<br>**Review period:** 1996 to March 2013<br>**Review purpose:** To evaluate the impact of prehabilitation on physical functional status, health care utilisation, quality of life, and pain after surgery<br>**Included study designs:** RCTs<br>**Included outcome measures:** | **Number of included studies:** 17<br>**Key characteristics:** Of the 17 RCTs, 13 studies were conducted in the orthopaedic setting: TKA, THA, and spinal surgery. The TKA and THA studies were conducted in patients with osteoarthritis. The remaining studies were conducted in the following settings: colorectal surgery, cardiac for participants awaiting coronary artery bypass graft, and upper gastrointestinal/hepatobiliary surgery. Age and gender of included participants not stated | Not prioritised due to more recent systematic reviews providing similar outcomes |
|  | Objective functional status, healthcare utilisation, quality of life and postoperative pain |  |  |
| Chen et al.(2018) Is it necessary to perform prehabilitation exercise for patients undergoing total knee arthroplasty: meta-analysis of randomized controlled trials<br>Vol 46, No 1<br>(tandfonline.com)<br>p.36-43. | **Intervention Type:** Exercise<br>**Review period:** January 1990 to June 2017<br>**Review purpose:** To test whether it is necessary to perform prehabilitation exercise for patients undergoing total knee arthroplasty (TKA)<br>**Included study designs:** RCTs<br>**Included outcome measures:** length of hospital stay, functional outcomes (quadriceps strength and functional ability) Sit-to-stand and 6-min walk, Knee range of motion (ROM), extension and flexion, WOMAC function, pain and stiffness | **Number of included studies:** 16<br>**Key characteristics:** A total of 16 studies were included in the review representing 1,224 participants (612 patients in the prehabilitation group and 612 patients in the control group), of which 770 were female and 454 were males. The average age of participants in the included studies ranged from 60 – 72 years | Not prioritised due to no information on the screening process, and many studies overlap with already included systematic reviews |
| Chesham and Shanmugam. (2017) Does preoperative physiotherapy improve postoperative, patient-based outcomes in older adults who have undergone total knee arthroplasty? A systematic review *Physiotherapy Theory and Practice*, 33(1), pp.9-30. | **Intervention Type:** Exercise<br>**Review period:** April 2004 to April 2014<br>**Review purpose:** Systematically review whether preoperative physiotherapy improves postoperative, patient-based outcomes in older adults who have undergone total knee arthroplasty (TKA) and compare study interventions to best-practice guidelines<br>**Included study designs:** RCTs<br>**Included outcome measures:** Treatment adherence, OA severity and surgery, follow-up and attrition, analgesics | **Number of included studies:** 10<br>**Key characteristics:** Nine hundred and nine participants were included in the ten studies, ranging from 17 to 243 participants and this included 545 females (60%) and 364 males (40%). The mean age of participants was 67.3 years, ranging from 63 years to 73 years. Mean BMI was 31.2 kg/m<sup>2</sup>, ranging from a mean of 27.1 kg/m<sup>2</sup> to 35.7 kg/m<sup>2</sup>. According to the BMI Classification, participants in three studies were classified as overweight, whereas participants in seven studies were classified as obese | Not prioritised as it did not meet our criteria for systematic reviews (screening and data extraction were only conducted by one reviewer) |
| Dennis et al. (2020) Effects of presurgical interventions on chronic pain after total knee replacement: a systematic review and meta-analysis of randomised controlled trials<br>BMJ Open 10(1), p.e033248. | **Intervention Type:** Exercise<br>**Review period:** Inception of databases to December 2018<br>**Review purpose:** To evaluate the effectiveness of presurgical interventions in preventing chronic pain after total knee replacement (TKR)<br>**Included study designs:** RCTs<br>**Included outcome measures:** Pain at 6 months or longer; adverse events | **Number of included studies:** 8<br>**Key characteristics:** Ten articles reporting data from eight RCTs were included in the review. These eight RCTs included data from 960 participants randomised to nine eligible comparisons. Five studies exclusively recruited participants undergoing total knee replacement (TKR) for osteoarthritis; one permitted those with rheumatoid arthritis; and two included participants undergoing either hip or knee replacement for osteoarthritis, with long-term data reported separately or in aggregate. Mean age of participants ranged from 63 to 70 across studies. Gender, where reported, varied greatly, with studies recruiting 17% to 60% women | Not prioritised as overlap of studies with Wang (2021) and some included studies from 1996 |
| Drudi et al. (2019) Preoperative Exercise Rehabilitation in Cardiac and Vascular Interventions<br>*journal of surgical research*, 237, pp.3-11. | **Intervention Type:** Exercise<br>**Review period:** Inception of databases to October 2016<br>**Review purpose:** To describe the current state of knowledge of the effects of preoperative exercise training (prehabilitation) on perioperative clinical outcomes in patients undergoing cardiac and vascular interventions<br>**Included study designs:** Randomised and non-randomised controlled trials<br>**Included outcome measures:** Length of stay, postoperative complications and quality of life | **Number of included studies:** 9<br>**Key characteristics:** Five RCTs and four Non-RCTs. The nine included studies were stratified by cardiac (n = 7) and vascular (n = 2) procedures. Among the cardiac procedures, the average age of participants ranged from 52 to 86 yrs. Most of the included studies consisted mainly of male participants ranging from 60% to 93%. The distribution of male and female participants was only balanced in one study. Among the vascular procedures, the average age of participants ranged from 65 to 73 yrs. The included studies consisted mainly of male participants ranging from 66% to 87% | Not prioritised due to study design and overlap of topics with other systematic reviews |
| Gill and McBurney. (2013) Does Exercise Reduce Pain and Improve | **Intervention type:** Exercise<br>**Review period:** up to July 2012 | **Number of included studies:** 18<br>**Key Characteristics:** A total of 18 studies were included in the review representing 1,028 | Not prioritised as It did not meet our criteria for a systematic review |
| Physical Function Before Hip or Knee Replacement Surgery? A Systematic Review and Meta-Analysis of Randomised Controlled Trials<br>*Archives of physical medicine and rehabilitation*, 94(1), pp.164-176. | **Review purpose:** To investigate the preoperative effects of exercise-based interventions on pain and physical function for people awaiting joint replacement surgery of the hip or knee.<br>**Included study designs:** Randomised or quasi-randomised control trials<br>**Included outcome measures:** pain and physical function including self-reported function, walking speed, and muscle strength | participants. Participants were awaiting knee replacement in 12 studies and hip replacement in 7 studies. One study included participants awaiting either hip or knee replacement. With the exception of 1 study where participants were noticeably younger, participants were typically older adults with average ages of between 60 and 80 years. Most studies had more women than men, and 1 study had only women | (screening not undertaken by multiple reviewers) |
| Gomes Neto et al. (2017). Pre- and postoperative inspiratory muscle training in patients undergoing cardiac surgery: systematic review and meta-analysis *Clinical rehabilitation*, 31(4) pp.454-464. | **Intervention Type:** Exercise (IMT)<br>**Review period:** 1950 to June 2015<br>**Review purpose:** To determine the effects of pre- and postoperative inspiratory muscle training on length of postoperative hospital stay and pulmonary function in patients undergoing cardiac surgery<br>**Included study designs:** RCTs<br>**Included outcome measures:** Postoperative complications, length of stay, Quality of life, inspiratory pressure, ventilation time, values for the six-minute walk test and anxiety/depression | **Number of included studies:** 8<br>**Key characteristics:** Four of the eight included studies investigated preoperative inspiratory muscle training (416 patients), three investigated postoperative inspiratory muscle training (115 patients) and one study investigated pre- and postoperative inspiratory muscle training (43 patients).<br>For studies of preoperative inspiratory muscle training, the final sample ranged from 26 to 276, and mean age of participants ranged from 60 to 70 years. Four studies included patients of both genders, but there was a predominance of male. In the studies of postoperative inspiratory muscle training, the final sample ranged from 30 to 47 and the mean age of participants ranged from 56 to 62 years. Three studies included patients of both genders, but there was a predominance of male. In one study that investigated the effects of pre- and postoperative inspiratory muscle training, the mean age of participants was of 60 years and included patients of both genders, but there was a predominance of male | Not prioritised due to overlap with other systematic reviews and a few included studies being conducted in non-OECD countries |
| Halloway et al. (2015) Prehabilitation interventions for older adults: an integrative review. *Western journal of nursing research*, 37(1), pp.103-123. | **Intervention type:** Exercise<br>**Review period:** up to January 2014<br>**Review purpose:** To examine the effect of prehabilitation interventions on physical activity behaviour and dimensions of physical fitness in older adults<br>**Included study designs:** RCTs<br>**Included outcome measures:** Physical activity and physical-fitness measures | **Number of included studies:** 7<br>**Key Characteristics:** Sample sizes ranged from 20 to 243. All papers focused on prehabilitation interventions for those undergoing total joint arthroplasty, either of the knee (n= 5), hip (n= 1) or either knee or hip (n= 1). The mean age of the participants from all seven studies was 69 years. For five studies, the proportion of women to men was similar whereas two studies had predominately female samples | Not prioritised as it did not meet our criteria for a systematic review (no quality appraisal undertaken) |
| Hoogeboom et al. (2012) Therapeutic Validity and Effectiveness of Preoperative Exercise on Functional Recovery after Joint Replacement: A Systematic Review and Meta-Analysis *PloS one*, 7(5), p.e38031. | **Intervention Type:** Exercise<br>**Review period:** up to January 2012<br>**Review purpose:** The aim of our study was threefold. First, we developed a rating scale to assess the therapeutic validity of therapeutic exercise programmes. Second, we assessed the therapeutic validity of preoperative therapeutic exercise programmes in patients awaiting elective, primary THR or TKR, and, finally, we assessed the association between therapeutic validity and the effect of the interventions on postoperative functional recovery<br>**Included study designs:** Randomised control trials or quasi-randomised control trials<br>**Included outcome measures:** In-hospital functional recovery, short-term observed functional recovery, short-term self-reported functional recovery | **Number of included studies:** 12<br>**Key characteristics:** A total of 12 articles comprising 11 RCTs and one quasi-randomised controlled trial met the eligibility criteria. One study presented data for both THR and TKR, eight interventions focused on TKR and five interventions on THR. These 12 primary studies included a total of 737 patients (55% women), with a mean (SD) age of 66 years and a Body Mass Index (BMI) of 31 | Not prioritised based on the year searches were conducted (2012) and the outcomes were covered by more recent systematic reviews |
| Hulzebos et al. (2012)<br>Preoperative physical therapy for elective cardiac surgery patients<br>*Cochrane database of systematic reviews* (11) | **Intervention Type:** Exercise<br>**Review period:** 1966 to Dec 2011<br>**Review purpose:** To determine if preoperative physical therapy with an exercise component can prevent postoperative pulmonary complications in cardiac surgery patients, and to evaluate which type of patient benefits and which type of physical therapy is most effective<br>**Included study designs:** Randomised control trials<br>**Included outcome measures:** Postop pulmonary complications grade 2, 3, 4; Postoperative pulmonary complications > grade 2, Postoperative all-cause mortality, adverse outcomes, Length of postoperative hospital stay, Physical function measures, Postoperative mortality from respiratory causes, Health-related quality of life | **Number of included studies:** 8<br>**Key characteristics:** Eight RCTs with 856 patients were included. Three studies used a mixed intervention (including either aerobic exercises or breathing exercises); and five studies used inspiratory muscle training. All patient characteristics that were reported on were evenly distributed per treatment group, except in one study (Ferreira 2009) where the experimental group had fewer males (60% versus 87% in the control group; $P < 0.01$). In one other study the median age in the experimental group was lower, although not significantly so, compared to the median age of controls (62.5 versus 68 years; $P = 0.06$) | Not prioritised based on the year searches were conducted and the outcomes were covered by more recent systematic reviews |
| Kwok et al. (2015)<br>Does Pre-Operative Physiotherapy Improve Outcomes in Primary Total Knee Arthroplasty? — A Systematic Review<br>*The Journal of arthroplasty*, 30(9), pp.1657-1663. | **Intervention type:** Exercise<br>**Review period:** 1946 to June 2014<br>**Review purpose:** To evaluate the evidence on whether preoperative physiotherapy improves patient outcomes following TKA<br>**Included study designs:** RCTs or quasi-randomised trials<br>**Included outcome measures:** postoperative WOMAC scores, lower limb strength, pain, range of movement and hospital length of stay | **Number of included studies:** 11<br>**Key Characteristics:** A total of 11 RCTs were included, representing 791 participants. The average ages reported for included studies ranged from 59 to 73 however for one study no information on age was provided. The average percentage of females in the included studies ranged from 20% to 89% however one of the included studies did not report the gender distribution. Most interventions involved multi-modal physiotherapy, encompassing a combination of different types of exercises – warm-up, aerobic exercise, resistance training, flexibility training, proprioceptive training and practicing of functional tasks – with or without patient education. However, the components of exercise programmes were very<br>variable in different studies, with one utilising proprioceptive training only and another four which did not include aerobic exercise training | Not prioritised as it did not meet our criteria for systematic reviews (study selection methods not described) |
| Lemanu et al. (2013)<br>Effect of Preoperative Exercise on Cardiorespiratory Function and Recovery After Surgery: a Systematic Review<br>*World journal of surgery*, 37(4), pp.711-720. | **Intervention Type:** Exercise<br>**Review period:** Inception of databases to December 2011<br>**Review purpose:** To investigate the extent to which preoperative conditioning (PREHAB) improves physiologic function and whether it correlates with improved recovery after major surgery<br>**Included study designs:** RCTs<br>**Included outcome measures:** cardiorespiratory physiological function, clinical outcomes (including length of hospital stay and rates of postoperative complications), and measures of changes in functional capacity (physical and psychological) | **Number of included studies:** 8<br>**Key characteristics:** The included studies involved a total of 868 patients, 442 of whom were randomized to a preoperative exercise program and 426 to either a control or “sham” intervention group. The patients were elderly (mean age >60 years), Gender distribution of included participants not stated | Not prioritised based on the year searches were conducted (2011) and the outcomes were covered by more recent systematic reviews |
| Mans et al. (2015)<br>Postoperative outcomes following preoperative inspiratory muscle training in patients undergoing cardiothoracic or upper abdominal surgery: a systematic review and meta analysis<br>*Clinical rehabilitation*, 29(5), pp.426-438. | **Intervention Type:** Exercise (IMT)<br>**Review period:** inception of databases to December 2013<br>**Review purpose:** To evaluate whether preoperative inspiratory muscle training is effective in preventing postoperative pulmonary complications and reducing length of hospital stay in people undergoing cardiothoracic or upper abdominal surgery<br>**Included study designs:** RCTs<br>**Included outcome measures:** Postoperative pulmonary complications, length of stay, maximal inspiratory pressure, maximal expiratory<br>pressure, inspiratory muscle endurance, forced expiratory volume in one second, forced vital capacity, exercise tolerance, walking distance, risk of requiring greater than 24 hours of mechanical ventilation postoperatively, patient satisfaction and adverse events | **Number of included studies:** 8<br>**Key characteristics:** Eight RCTs provided data on 295 participants, with individual sample sizes ranging from 20 to 84 participants. Four studies (n = 180) investigated participants undergoing cardiac surgery, one study (n = 23) investigated those undergoing thoracic surgery, and three studies (n = 92) investigated participants undergoing upper abdominal surgery. The mean age of participants ranged from 35 to 71 years. In total, 53% of participants were male | Not prioritised due to overlap with Katsura (2015) |
| Marmelo et al. (2018) The impact of prehabilitation on post-surgical complications in patients undergoing non-urgent cardiovascular surgical intervention: Systematic review and meta-analysis *European journal of preventive cardiology*, 25(4), pp.404-417. | **Intervention Type:** Exercise (IMT)<br>**Review period:** 2000 to November 2016<br>**Review purpose:** Evaluating the ability of prehabilitation to prevent post-surgical complications in cardiac patients<br>**Included study designs:** RCTs, non-randomised control trials and quasi-randomised control trials<br>**Included outcome measures:** Postoperative complications, length of stay, Quality of life, inspiratory pressure, ventilation time, values for the six-minute walk test and anxiety/depression | **Number of included studies:** 8<br>**Key characteristics:** Of the eight studies selected (6 RCTs, 1 NON-RCT and 1 Quasi-RCT), four had samples with less than 50 participants, one had a sample size with 50 to 100 participants, whereas three studies had samples of more than 100 participants in a total of 947 individuals. 79.8% of study participants were males and 20.1% were females, with a mean age of $63.1 \pm 5.1$ years (range between 52.8 years and 70.5 years) | Not prioritised due overlap of included studies in Rodrigues (2021). Similar processes with Rodrigues (MA) and a few studies are from non-OECD countries |
| Palmer et al. (2020) Presurgery exercise-based conditioning interventions (prehabilitation) in adults undergoing lower limb surgery for peripheral arterial disease *Cochrane Database of Systematic Reviews*, (9). | **Intervention Type:** Exercise<br>**Review period:** up to September 2019<br>**Review purpose:** To assess the effectiveness of prehabilitation (preoperative exercise, either alone or in combination with nutritional or psychological interventions, or both) on postoperative outcomes in adults with PAD undergoing open lower limb surgery<br>**Included study designs:** RCTs | **Number of included studies:** 0<br>**Key characteristics:** No RCTs were identified that met the inclusion criteria | Not prioritised as it did not meet our criteria for a systematic review no studies were identified that met the inclusion criteria |
|  | **Included outcome measures:** Postoperative complications, mortality and readmission to hospital within 30 days of the surgical procedure |  |  |
| Pouwels et al. (2015). Beneficial Effects of Pre-operative Exercise Therapy in Patients with an Abdominal Aortic Aneurysm: A Systematic Review *European Journal of Vascular and Endovascular Surgery*, 49(1), pp.66-76. | **Intervention Type:** Exercise<br>**Review period:** Inception of databases to May 2014<br>**Review purpose:** This systematic review focuses on the possible effects of preoperative exercise therapy (PET) in patients with abdominal aortic aneurysm (AAA) on postoperative complications, aerobic capacity, physical fitness, and recovery<br>**Included study designs:** RCTs or prospective cohort trials<br>**Included outcome measures:** Patient satisfaction, respiratory function, postoperative complications, physical fitness parameters and compliance | **Number of included studies:** 5<br>**Key characteristics:** Four RCTs, one prospective cohort study representing a total 147 participants (116 males and 31 females). The average age of participants in the included studies ranged from 59 – 75 years. Three of the included studies investigated an exercise program in patients with an abdominal aortic aneurysm (AAA) (without indication for surgical repair with a duration ranging from 6 weeks to 12 months. One studied a PET program for 6 consecutive weeks in patients awaiting elective infrarenal AAA repair. One studied an inspiratory muscle training program for at least 2 weeks before surgery in patients awaiting elective AAA repair | Not prioritised as overlap of included studies with other systematic reviews |
| Santa Mina et al. (2014) Effect of total-body prehabilitation on postoperative outcomes: a systematic review and meta-analysis. *Physiotherapy* 100(3), pp.196-207. | **Intervention Type:** Exercise<br>**Review period:** 1950 to 2011<br>**Review purpose:** To systematically review the evidence of preoperative exercise, known as 'prehabilitation', on peri- and postoperative outcomes in adult surgical populations<br>**Included study designs:** RCTs, non-RCTs, single-group trials, prospective case-control, randomized case study<br>**Included outcome measures:** Preoperative physical function, postoperative health related quality of life, postoperative pain, | **Number of included studies:** 21<br>**Key characteristics:** Studies were published between 1996 and 2011. Of the 21 included trials, 17 were RCTs with one non-RCT, one single-group trial (with secondary outcomes published separately), one prospective case-control study using historical control subjects, and one randomised case study design. In total, 1371 participants were included in these 21 trials. Thirteen studies assessed preoperative exercise in orthopaedic populations, and eight studies examined preoperative exercise in patients undergoing visceral organ surgery. The median intervention duration was 6 weeks (range 1 to 8 | Not prioritised based on the year searches were conducted (2011) and the outcomes were covered by more recent systematic reviews |
|  | musculoskeletal and functional task performance, aerobic fitness, length of stay, healthcare utilisation, per-operative complications and adverse events | weeks). Age and gender distribution of included participants not stated |  |
| Shoemaker et al. (2013) Preoperative Exercise in Individuals undergoing total knee arthroplasty 29(1), pp.2-16. | **Intervention Type:** Exercise<br>**Review period:** up to April 2012<br>**Review purpose:** The purpose of this article was to systematically review all studies related to the efficacy of preoperative exercise for patients undergoing total knee arthroplasty and to consider issues not previously addressed by previous systematic reviews<br>**Included study designs:** RCTs, case series, case report, retrospective cohort study<br>**Included outcome measures:** Impairment-based outcome measures (range of motion, strength in subjects, knee extension, pain, knee, proprioception, quadriceps muscle cross-section area); Activity-based outcome measures; participation-based outcome measures (WOMAC, SF-36 score, HSS knee rating score, Knee society score); Health care utilisation outcomes (Length of hospital stay, need for outpatient physical therapy, amount of assistance required on the first postoperative, overall cost, discharge setting) | **Number of included studies:** 17<br>**Key characteristics:** A total of 17 studies were included in the review (13 RCTs, one retrospective cohort study, two prospective case series and one case report), representing 1,015 participants (age and gender distribution not stated for all included studies). Five of 17 studies measured range of motion, nine studies measured strength in subjects, three studies measured pain on a visual analogue scale | Not prioritised based on the year searches were conducted (2012) and the outcomes were covered by more recent systematic reviews |
| Tew et al. (2022) Preoperative exercise training for adults undergoing elective major vascular surgery: A systematic review *PloS one*, 17(1), p.e0263090. | **Intervention Type:** Exercise<br>**Review period:** January 2008 to April 2021<br>**Review purpose:** To assess the benefits and harms of preoperative exercise training in adults undergoing elective vascular surgery<br>**Included study designs:** RCTs | **Number of included studies:** 3<br>**Key characteristics:** A total of 197 participants were included in these studies, with study sample sizes ranging from 20 to 124. Group mean ages ranged from 59 to 75 years across the studies. In two studies participants were mostly male, and in the third study they were mostly female. All three studies involved people undergoing abdominal | Not prioritised as overlap of studies with Fenton (2021) who also conducted a meta-analysis |
|  | **Included outcome measures:** Mortality, complications, HRQOL, adverse events, adherence to programme | aortic aneurysm (AAA) repair; a mixture of open and endovascular repairs in two studies, and unspecified in the other |  |
| Thybo Karanfil and Møller. (2018)<br>Preoperative inspiratory muscle training prevents pulmonary complications after cardiac surgery—a systematic review. *Dan Med J*, 65(3), p.A5450. | **Intervention Type:** Exercise (IMT)<br>**Review period:** inception of databases to September 2017<br>**Review purpose:** The aim of this systematic review was to determine if preoperative inspiratory muscle training could prevent the development of pneumonia and atelectasis in patients undergoing coronary artery bypass grafting (CABG) or heart valve surgery<br>**Included study designs:** RCTs<br>**Included outcome measures:** Incidence of atelectasis and incidence of pneumonia, pleural effusion | **Number of included studies:** 5<br>**Key characteristics:** The total number of patients in the analysis was 451. Median study size was 32 (range: 26-276), Male gender accounted for the main part of the subjects of each trial with a median study size of 22 (range: 13-215), whereas female gender presented a median study size and range of 13 (range: 8-61). Four studies had only CABG patients in their population, and one study investigated a population comprising both CABG patients and patients undergoing heart valve surgery | Not prioritised due to all included studies being captured by Rodrigues 2021 which is a similar more up to date SR |
| Valkenet et al. (2011)<br>The effects of preoperative exercise therapy on postoperative outcome: a systematic review<br>*Clinical rehabilitation*, 25(2), pp.99-111. | **Intervention Type:** Exercise<br>**Review period:** Inception of databases to January 2010<br>**Review purpose:** To summarize the current evidence on the effects of preoperative exercise therapy in patients awaiting invasive surgery on postoperative complication rate and length of hospital stay<br>**Included study designs:** Controlled trials<br>**Included outcome measures:** Postoperative pulmonary complications, length of hospital stay, postoperative complication rates | **Number of included studies:** 12<br>**Key characteristics:** A total of 12 studies were included in the review representing 1,245 participants (870 males, 375 females). The average age of participants in the included studies ranged from 45 – 71 years. Seven studies included patients awaiting joint replacement (total knee arthroplasty or total hip arthroplasty). Four studies described the effects of preoperative exercise therapy on candidates for coronary artery bypass graft surgery. One study concerned patients with an abdominal aortic aneurysm awaiting abdominal surgery | Not prioritised based on the year searches were conducted (2010) and the outcomes were covered by more recent systematic reviews |
| Wee and Choong. (2020) A systematic review of the impact of preoperative exercise for patients with abdominal aortic aneurysm *Journal of Vascular Surgery*, 71(6), pp.2123-2131. | **Intervention Type:** Exercise<br>**Review period:** Inception of databases to May 2018<br>**Review purpose:** This review provides comprehensive coverage of both physical fitness parameters and postoperative clinical outcomes. We hypothesize that preoperative exercise will improve postoperative outcomes and physical fitness parameters in abdominal aortic aneurysm (AAA) patients<br>**Included study designs:** RCTs and retrospective cohort study<br>**Included outcome measures:** Compliance and patient satisfaction, physical fitness capacity, postoperative complications, and other clinical outcomes (preoperative respiratory function, length of hospital stay) | **Number of included studies:** 7<br>**Key characteristics:** Six RCTs and one Retrospective Cohort study representing 371 participants (322 males, 49 females). Ages ranged from 59 to 80 years. Three studies assessed the impact of an exercise program in AAA patients who were not waiting for surgical repair. The remaining four studies investigated an exercise program in abdominal aortic aneurysm (AAA) patients who were waiting for surgical repair | Not prioritised as overlap of studies with Fenton (2021) who also conducted a meta-analysis |
| Educational interventions |  |  |  |
| Citation | Review details | Included studies | Comments /notes |
| Aydin et al. (2015) No major effects of preoperative education in patients undergoing hip or knee replacement--a systematic review. *Dan Med J*, 62(7), p.A5106. | **Intervention type:** Educational<br>**Review period:** up to October 2014<br>**Review purpose:** To determine whether the literature supports a positive effect of PPE on postoperative outcomes including anxiety, pain, length of hospital stay (LOS), patient satisfaction, postoperative complications, mobility, and expectation<br>**Included study designs:** RCTs | **Number of included studies:** 12<br>**Key Characteristics:** A total of 1,567 participants were included from 12 studies (619 males and 948 females). Six studies involved patients undergoing THA, five studies involved both THA and TKA, and one study TKA only. Preoperative education was given within six weeks (range: one day - six weeks) of arthroplasty surgery. The methods used to provide the preoperative education varied: booklets, seminars, videotapes. The mean age of participants in the included studies ranged from 62.4 to 71.9 years, however for one study a mean age of participants was not stated | Not prioritised as it did not meet our criteria for a systematic review (no quality appraisal undertaken) |
|  | **Included outcome measures:** Length of stay, anxiety, pain, patient satisfaction, patient expectations and postoperative mobility |  |  |
| McDonald et al. (2014) Preoperative education for hip or knee replacement. Cochrane database of systematic reviews, (5). | **Intervention Type:** Educational<br>**Review period:** 1872 to May 2013<br>**Review purpose:** To determine whether preoperative education in people undergoing total hip replacement or total knee replacement improves postoperative outcomes with respect to pain, function, health-related quality of life, anxiety, length of hospital stay and the incidence of adverse events (e.g. deep vein thrombosis)<br>**Included study designs:** RCTs and quasi-randomised control trials<br>**Included outcome measures:** Postoperative pain, function, length of hospital stay, adverse events, quality of life, preoperative anxiety, postoperative anxiety, postoperative range of motion and postoperative mobility | **Number of included studies:** 18<br>**Key characteristics:**<br>A total of 18 trials (17 of which were RCTs, and one was a Quasi-RCT) representing 1,463 participants were included in the review. Of the 18 trials, 13 involved people undergoing hip replacement, three involved people undergoing knee replacement and two included both people with hip and knee replacements. Most participants were women (59%), and the mean age of participants was within the range of 58 to 73 years | Not prioritised based on the year searches (2013) were conducted and the outcomes were covered by more recent systematic reviews |
| Ronco et al. (2012) Patient education outcomes in surgery: a systematic review from 2004 to 2010 International journal of evidence-based healthcare, 10(4), pp.309-323. | **Intervention Type:** Educational<br>**Review period:** January 2004 to April 2010<br>**Review purpose:** To describe preoperative educational interventions (including content and delivery time) and postoperative outcomes as considered in studies evaluating the effectiveness for patients undergoing major surgery published from 2004 to 2010<br>**Included study designs:** RCTs and clinical trials | **Number of included studies:** 19<br>**Key characteristics:** A total of 19 studies involving 3,944 patients were included. Of these, 12 were RCTs and seven were clinical trials (with control and experimental groups not randomised). The mean age of participants was 59.7 years, and 61.7% of participants were male. Of the studies included, seven addressed orthopaedic surgery and involved a total of 2,072 patients (52.5% of total), six cardiac surgery (1,202 patients, 30.4%), three major surgery not specified (465 patients, 11.8%), | Not prioritised due to year searches conducted (2010) and more recent SRs |
|  | **Included outcome measures:** Experiential outcomes (postop anxiety, satisfaction, preoperative anxiety, preoperative depression, postoperative depression, quality of life, health status, self-efficacy, and emotional wellbeing); Cognitive outcomes (objective knowledge, subjective knowledge, and patient's evaluation of quality and satisfaction of the education received); Biophysiological outcomes (postoperative pain intensity, perceived control of pain, blood pressure and heart pulse rate, amount of morphine administered, and postoperative agitation/confusion); Financial outcomes (length of stay, need for additional postop treatment, related costs); Functional outcomes (daily living activities compromised by surgery, or by pain); Ethical outcomes (patient awareness of legal rights); Complications (drug-related, functional postoperative complications) | two abdominal surgery (135 patients, 3.4%) and one chest surgery (75 patients, 1.9%) |  |
| **Weight loss interventions** |  |  |  |
| Lui et al. (2015)<br>Effect of non-surgical, non-pharmacological weight loss interventions in patients who are obese prior to hip and knee arthroplasty surgery: a rapid review<br>*Systematic reviews*, 4(1), pp.1-8. | **Intervention type:** Weight Loss<br>**Review period:** January 1990 to February 2015<br>**Review purpose:** To examine the effects of short-term non-pharmacological and non-surgical weight loss interventions in adults in the year prior to total hip arthroplasty (THA) and total knee arthroplasty (TKA) on surgical and patient outcomes, and adverse events<br>**Included study designs:** Retrospective cohort studies<br>**Included outcome measures:** Surgical site infection (deep and superficial), 90-day readmission, adverse events | **Number of included studies:** 2<br>**Key characteristics:** Two Cohort studies were included in the review representing 24,459 participants (6,620 total hip arthroplasty patients and 17,839 total knee arthroplasty). In one study 43.9% of participants were <65 years of age and in the other 46.6% of participants were <65 years of age. However, the age and gender distribution of all included participants were not stated | Not prioritised due to study designs and no intervention details provided |

**Appendix 4:** Summary of ongoing systematic reviews

| Ongoing reviews |  |  |  |
| --- | --- | --- | --- |
| Citation<br>(Country) | Review details | Inclusion/exclusions criteria | Reviewer comments |
| Rombey T, Eckhardt H, and Quentin W. (2020). Cost-effectiveness of prehabilitation prior to elective surgery compared to usual preoperative care: protocol for a systematic review of economic evaluations *BMJ open*, 10(12), p.e040262. | **Intervention type:** Prehabilitation - exercise<br>**Review period:** Anticipated completion date was 31 March 2021<br>**Review purpose:** The aim of this systematic review is to investigate the cost-effectiveness of prehabilitation programmes for patients awaiting elective surgery compared with usual preoperative care | **Inclusion:**<br>**Population** – Patients from any surgery undergoing elective surgery<br>**Intervention** – A preoperative prehabilitation programme (any setting), defined as a (set of) intervention(s) aimed to optimise functioning and reduce disability in individuals awaiting surgery. The intervention(s) had to include at least one component of physiotherapy or occupational therapy and at least one in-person meeting between the patient(s) and healthcare professional(s). The programme's overall duration and the duration and frequency of individual sessions had to be sufficiently long to have an effect given the patients fully adhered to it.<br>**Control** – Usual preoperative care as defined by the study authors, that is, the routine care that patients with a given condition receive in the respective hospital (extended only by the baseline measurements performed as part of the trial)<br>**Outcome** – Clinical effectiveness and costs, any time frame for follow-up<br>**Study type** – Full (i.e., cost-benefit, cost-effectiveness and cost-utility analyses) or partial economic evaluations (i.e., cost-minimisation analysis), trial based (any design) or model based. Studies will be included regardless of their cost perspective, publication year, language and status (i.e., full article, protocol/registration record, conference abstract)<br>**Exclusion:**<br>**Population** – Patients undergoing emergency surgery or non-surgical treatments (e.g., chemotherapy)<br>**Intervention** – Purely medical/nutritional interventions (e.g., an extra dose of a specific drug/nutritional supplements), an intervention consisting of prehabilitation combined with additional postoperative rehabilitation, a prehabilitation programme not containing any physiotherapy or occupational therapy components (e.g., cognitive-behavioural therapy or health counselling/education only), a prehabilitation programme without any in-person meetings (e.g., purely web/app based).<br>**Control** – Another prehabilitation intervention<br>**Outcome** – Clinical effectiveness only | Review authors contacted on 01/03/22 and full publication is anticipated at the end of March 2022 |
|  |  | **Study type** – Systematic reviews, mere cost analyses (i.e., studies that simply calculated the costs of the intervention but did not compare it to the costs of for the control treatment), commentaries/letters, animal studies |  |
| Wright S, Wiechula R, and McLiesh P. (2016). The effectiveness of prehabilitation for adults having elective surgery: a systematic review protocol. *JBIEvidence Synthesis*, 14(2), pp.78-92. | **Intervention type:** Prehabilitation – multicomponent<br>**Review period:** No anticipated completion date given<br>**Review purpose:** The objective of this review is to identify the effectiveness of prehabilitation on postoperative outcomes of adults undergoing elective surgery | **Inclusion:**<br>**Types of participants** – Adult patients, aged 18 years or over, who are undertaking elective surgery in hospitals, including day surgery.<br>**Types of interventions** – Any prehabilitation programs and strategies that support early intervention. These include interventions such as early physical assessment, nutritional counselling, protein supplementation, weight loss initiatives, patient education, psychosocial support, self-instructed physical program and/or professional intervention for improved physical function, including physiotherapy and comorbidity resolve. These intervention strategies will be compared with the “usual care” patients receive prior to surgery. Only studies with multimodal prehabilitation interventions would be selected<br>**Types of outcomes** – **Primary outcomes:** postoperative complications and patient readmission, length of stay, length of rehabilitation required and HRQOL. **Secondary outcomes:** delay in surgery, admission into respite or residential care. Health Related Quality of Life outcomes will be measured using the MOS (Medical Outcomes Study) 36-Item Short-Form Health Survey (SF-36)<br>**Types of studies** – This review will consider experimental and epidemiological study designs including randomized controlled trials, non-randomized controlled trials, quasi-experimental studies, before and after studies, prospective and retrospective cohort studies, case control studies and analytical cross sectional studies for inclusion. As studies are likely to vary considerably both in terms of populations, interventions and outcomes, the review will also consider descriptive epidemiological study designs including case series, individual case reports and descriptive cross sectional studies where higher level studies are not identified for those criteria<br>**Exclusion:** Studies that include interventions initiated within one month prior to surgery will be excluded. This is to distinguish between prehabilitation interventions and short-term interventions such as preoperative assessment clinics. Interventions addressing exercise therapy as the only component will not be included as this has been covered in previous reviews | Review authors contacted on 01/03/22, still awaiting response |
| Tait et al. ( 2020) A Network Meta-analysis assessing the Impact of | **Intervention type:** Exercise<br>**Review period:** Anticipated completion date 31 Dec 2020 | **Inclusion:**<br>**Types of study** – All prospective trials, including randomised controlled trials and cohort studies/prospective observational studies. | Review authors contacted on 01/03/22, still |
| Preoperative Exercise on Fitness and Clinical Postoperative outcomes.<br>PROSPERO<br>CRD42020205078 | **Review purpose:** What impact do different modalities of preoperative exercise have on patients' fitness, and how does this effect postoperative outcomes? | **Condition or domain being studies** – Prehabilitation interventions before elective surgery (including orthopaedic, abdominal and thoracic surgery) for a range of malignant (oncological surgery) and benign (including bariatric surgery) conditions. Prehabilitation had to include aerobic exercise, strength/resistance training or inspiratory muscle training or a combination of these. Additional interventions including education, nutritional and psychological interventions will be summarised with the above interventions.<br>**Population** – Adult (18 yrs. and older) patients undergoing elective surgery in prospective studies, comparing an exercise intervention to standard care or another exercise intervention. Studies where prehabilitation was administered after neoadjuvant chemotherapy and/or radiotherapy (NACRT) and before surgery were included.<br>**Interventions** – Aerobic Exercise: High Intensity aerobic training, Moderate intensity training, Low intensity training (including 'green' prescription), Strength/resistance training, Inspiratory muscle training (IMT)/respiratory training/chest physiotherapy, Aerobic exercise (Moderate and high intensity) and Strength/resistance training, Low intensity Aerobic exercise and Low intensity strength training, IMT and aerobic exercise (Moderate and high intensity), Aerobic (Moderate and high intensity) and Strength/resistance training and IMT. Control group: Standard Care. Additional interventions including education, nutritional and psychological interventions will be included with the above interventions/exposures<br>**Outcomes** – Change in fitness assessed using: Peak Oxygen Consumption (VO<sub>2</sub>Peak/Max) 6-Minute Walk Test. Incidence of postoperative complications. All complications as reported by the authors<br>**Exclusion:**<br>Studies where neoadjuvant chemotherapy and/or radiotherapy (NACRT) was being simultaneously given were excluded. When a preoperative intervention was targeted to treat a specific pathology before a specific operation, the study will be excluded | awaiting response |
| Qin C. et al. (2018)<br>Available from:<br>Effects of preoperative exercise training on postoperative complications of patients undergoing abdominal surgery - | **Intervention type:** Exercise<br>**Review period:** Anticipated completion date 17 October 2018<br>**Review purpose:** To explore whether preoperative exercise training could improve the | **Inclusion:**<br>**Types of study** – Only randomized controlled studies will be included,<br>**Participants** – Patients undergoing abdominal surgery,<br>**Intervention** – Preoperative exercise training/ prehabilitation, Patients in intervention group performed preoperative exercise program, and Patients in control group did not,<br>**Outcomes** – Outcome of postoperative complications, additional outcomes include length of stay, mortality and function capacity<br>**Exclusion:** | Review authors contacted on 01/03/22, still awaiting response |
| a meta-analysis of randomized controlled trials.<br>PROSPERO<br>CRD42018110752 | postoperative outcomes of abdominal surgery, especially postoperative complications, including all complications, major complications and pulmonary complications | Studies did not comprise outcome of interest, Full-text articles not available, Cohort or case-control studies, Patients who had undergone other kinds of surgery other than abdominal surgery, Studies in which patients in the control group performed different extents of preoperative exercise program, One-arm studies, Reanalysis of randomized controlled study |  |
| Hierro A. (2017)<br>The effectiveness of preoperative therapeutic exercise for recovery after hip or knee arthroplasty in patients with osteoarthritis<br>PROSPERO<br>CRD42017069044 | **Intervention type:** Exercise<br>**Review period:** Anticipated completed date 18 December 2017<br>**Review purpose:** What is the effectiveness of exercise after hip or knee arthroplasty? Does presurgical exercise improve the activities of daily living after surgery? | **Inclusion:**<br>**Types of study** – randomized controlled trials and prospective randomized trials, **Participants** – patients with osteoarthritis of the hip or knee on the waiting list for arthroplasty. The study sample must include participants aged 50 years or above. Osteoarthritis of the hip or knee.<br>**Intervention** – pre-surgical exercise. The effects of strength- or endurance-enhancing exercises on the activities of daily life in patients with osteoarthritis on the waiting list for arthroplasty, compared with normal care of the patient, in addition to improving the patient's range of movement<br>**Outcomes** – Changes in the activities of daily living, as measured using subjective scales for osteoarthritis (WOMAC), the hip (HOOS) and the knee (KOOS). Assessment will also be based on measurement using test objectives, such as the six minute walk test<br>**Exclusion:**<br>Participants under the age of 50 | Review authors contacted on 01/03/22, still awaiting response |
| Kunadharaju R. et al. (2021).<br>Effects of preoperative respiratory muscle training exercises on postoperative outcomes for patients undergoing thoracic surgery: a network meta-analysis<br>PROSPERO<br>CRD42021231315 | **Intervention type:** Exercise (respiratory muscle training)<br>**Review period:** Anticipated completion date 30 April 2021<br>**Review purpose:** What are the Effects of Preoperative Respiratory Muscle Training Exercises on Postoperative Outcomes for Patients Undergoing Thoracic Surgery? | **Inclusion:**<br>**Types of study** – RCTs, controlled clinical trials, pragmatic clinical trials, preoperative trial<br>Participants – Adult population who are undergoing Thoracic surgery - Coronary artery bypass surgery, Lung resection - Open or VATS, Esophageal surgeries, Aortic aneurysm repair, Open Aortic valve or Mitral Valve<br>**Surgery** – Thoracic surgery, cardiac surgery lung/esophageal. All thoracic surgery, **Intervention** – Exercise training, respiratory muscle training (breathing exercises) or both<br>**Exclusion:**<br>Studies will be excluded if they do not concern thoracic surgery, cardiac surgery lung/esophageal or thoracic surgery. If they do not include breathing exercises preoperatively, if they are the wrong study design, do not have measurable study data<br>or a control group. If it is a conference proceedings, duplicate, not in English or includes the wrong population | Review authors contacted on 01/03/22, still awaiting response |
| Jørgensen S, L. (2021)<br>The effectiveness of preoperative exercise performed to promote skeletal muscle hypertrophy and increase muscle strength prior to total joint replacement on postoperative functional outcome – A systematic Review and Meta-analysis of Randomized Controlled Trials<br>PROSPERO<br>CRD42021264796 | **Intervention type:**<br>Exercise<br>**Review period:**<br>Anticipated completion date is 31 August 2022<br>**Review purpose:**<br>To examine the effectiveness of preoperative exercise intervention performed to promote skeletal muscle hypertrophy and increase muscle strength prior to total knee replacement when assessed postoperatively for the following outcome measures; performance-based functional capacity, maximal lower limb muscle strength and patient-reported outcome on functional capacity, activities of daily living and pain 2-4 months postoperative, and 5-7 and 10-12 months postoperative. | **Inclusion:**<br>**Types of study** – RCTs<br>**Population** – people suffering from knee or hip osteoarthritis scheduled for a total joint replacement.<br>**Intervention** – Exercise paradigms designed and implemented to increase lower limb muscle strength and promote skeletal muscle hypertrophy. Specifically, this could be (i) resistance training where the exercise intensity is reported as a percentage of repetition maximum (RM) 1RM performed at least 2 times per week with or without blood flow restriction, or (ii) resistance training at an unspecific intensity performed to failure in at least one set per exercise performed at least 2 times per week<br>**Comparison** – usual care<br>**Outcome** – Performance-based measures of functional capacity at 2-4 months, 5-7 and 10-12 months after surgery, 30-seconds sit to stand, timed up and go, stair climbing, walking test | Review authors contacted on 01/03/22, responded saying they were close to submitting for publication (no dates given) |
| McCaffrey N, Higgins J, and La A. (2021)<br>A systematic review of economic evaluations of interventions for preoperative smoking cessation to prevent surgical complications | **Intervention type:** Smoking cessation<br>**Review period:**<br>Anticipated completion date 01 July 2022<br>**Review purpose:**<br>Which preoperative smoking cessation interventions are | **Inclusion:**<br>**Types of study** – economic evaluations<br>**Intervention** – all smoking cessation interventions<br>**Population** – smokers who are undergoing surgery<br>**Outcomes** – surgical complications, incremental cost-effectiveness ratio, Evaluations which report the costs and outcomes for at least two alternatives will be considered if the incremental cost-effectiveness ratio can be calculated manually<br>**Exclusion:** | Review authors contacted on 01/03/22, responded saying they were close to submitting for publication (expected to be |
| PROSPERO<br>CRD42021257740 | cost-effective for preventing surgical complications?<br>What are the costs and benefits of preoperative smoking cessation interventions for preventing surgical complications in different types of surgery? | **Types of study** – Health services costing studies, partial economic evaluations, editorials, comment or discussion papers, qualitative studies, reviews, case reports, case series, book chapters and conference articles | published July/Aug 2022) |
| Hayashi K. et al. (2020).<br>Impact of preoperative exercise on postoperative pain across the operation: A Systematic Review and Meta-Analysis of randomized controlled trials.<br>PROSPERO<br>CRD42020202701 | **Intervention type:** Exercise<br>**Review period:** Anticipated completion date 04 February 2021<br>**Review purpose:** Is there an impact of preoperative exercise on reduction of postoperative pain across the operation? | **Inclusion:**<br>**Types of study** - a full article with a randomized design comparing preoperative active exercise program versus no formal program will be included<br>**Population** – participants are waiting for every operation<br>**Outcome** – the outcome of interest is postoperative pain. The Likert scales of participant-reported pain, such as a Visual Analogue Scales (VAS), and a Numerical Rating Scales (NRS) will be used. If the Likert scales of pain will be not used, the following scales related pain will be used in sequence; a 36-item Short-Form Health Survey (SF-36) Bodily Pain subscale, West Haven-Yale Multidimensional Pain Inventory (WHYMPI), Western Ontario and McMaster Universities Osteoarthritis Index (WOMAC) pain subscale, Knee injury and Osteoarthritis Outcome Score (KOOS) pain subscale, and Hip Disability and Osteoarthritis Outcome Score (HOOS) pain subscale<br>**Exclusion:**<br>**Types of study** – reviews, editorials, case reports, and non-primary articles will be excluded<br>**Intervention** – intervention group participants receive not active exercise program, such as only passive stretching exercise, education, or psychological treatment. Intervention group participants receive preoperative program of pharmacological treatment, except for usual care. Intervention group participants receive intervention extended into the postoperative period. Control group receive preoperative or postoperative exercise program apart from usual care. Recruit the same patients with other articles | Review authors contacted on 01/03/22, responded saying the paper is under second review and hoping to be published soon (no dates given) |
| Punnoose A. et al. (2019)<br>Effectiveness of prehabilitation for patients undergoing orthopaedic surgery: | **Intervention type:**<br>Prehabilitation (exercises/physiotherapy, patient education, pain management and anxiety reduction strategies) | **Inclusion:**<br>**Types of study** – randomised controlled trials comparing prehabilitation (including multimodal prehabilitation interventions) to standard care<br>**Participants** – adult participants (>18years) undergoing an orthopaedic surgical procedure | Review authors contacted on 01/03/22, responded saying they were close to |
| protocol for a systematic review and meta-analysis<br>**BMJ open, 9(11), p.e031119** | **Review period:** Anticipated completion date not specified<br>**Review purpose:** The aim of this study is to evaluate the effectiveness of prehabilitation for patients undergoing orthopaedic surgery including day surgery procedures. It will also investigate the components of prehabilitation to understand optimum duration and frequency of programmes. | **Intervention** – prehabilitation or preoperative interventions including exercises/physiotherapy, patient education, pain management and anxiety reduction strategies<br>**Comparators** – Standard or usual care<br>**Outcomes** – Pain, muscle strength, function, health-related quality of life (HRQoL) and disease specific/ joint specific outcomes<br>**Publication** – As the researchers only began investigating the effectiveness of prehabilitation on postoperative outcomes since the turn of the millennium, only trials published between January 2000 to 25 March 2019 would be included<br>**Exclusion** – all non-English publications will be excluded (at full text stage) to avoid introducing risk of bias | submitting for publication (no dates given) |

## REFERENCES

Alshewaier, S., Yeowell, G., and Fatoye, F. (2017) The effectiveness of pre-operative exercise physiotherapy rehabilitation on the outcomes of treatment following anterior cruciate ligament injury: a systematic review. Clinical Rehabilitation, 31, 34–44.

Blasco, J.M., Hernandez-Guillen, D., Dominguez-Navarro, F., Acosta-Ballester, Y., Alakhdar-Mohmara, Y. and Roig-Casasus, S. (2021). Sensorimotor training prior total knee arthroplasty and effects on functional outcome: A systematic review and meta-analysis. Gait & Posture, 86, 83–93.

Burgess, L, C., Arundel, J., and Wainwright, T, W. (2019). The Effect of Preoperative Education on Psychological, Clinical and Economic Outcomes in Elective Spinal Surgery: A Systematic Review. Healthcare, 7, 48.

Carr, A., Smith, J. A., Camaradou, J. and Prieto-Alhambra, D. (2021). Growing backlog of planned surgery due to covid-19. BMJ, 372.

Centre for perioperative care (2021). Tackling the elective surgery backlog: Perioperative care solutions to the waiting list. Churchill House, 35 Red Lion Square London WC1R 4SG: Centre for Perioperative Care. Available at: https://www.cpoc.org.uk/sites/cpoc/files/documents/2021-09/CPOC-Perioperative-Care-Solutions-FINAL.pdf

Fenton, C., Tan, A.R., Abaraogu, U.O., and MCcaslin, J. E. (2021). Prehabilitation exercise therapy before elective abdominal aortic aneurysm repair. Cochrane Database of Systematic Reviews.

Gagliardi, A.R., Yip, C.Y., Irish, J., Wright, F.C., Rubin, B., Ross, H., Green. R., Abbey, S., McAndrews, M.P. and Stewart, D.E. (2021). The psychological burden of waiting for procedures and patient-centred strategies that could support the mental health of wait-listed patients and caregivers during the COVID-19 pandemic: A scoping review. Health Expectations, 24, 978–990.

Husted, R.S., Juhl, C., Troelsen, A., Thorborg, K., Kallemose, T., Rathleff, M.S. & Bandholm, T. (2020). The relationship between prescribed pre-operative knee-extensor exercise dosage and effect on knee-extensor strength prior to and following total knee arthroplasty: a systematic review and meta-regression analysis of randomized controlled trials. Osteoarthritis and Cartilage, 28, 1412–1426.

Katsura, M., Kuriyama, A., Takeshima, T., Fukuhara, S. and Furukawa, T.A. (2015). Preoperative inspiratory muscle training for postoperative pulmonary complications in adults undergoing cardiac and major abdominal surgery. Cochrane Database of Systematic Reviews.

Levy, N., Selwyn, D.A. and Lobo, D.N. (2021). Turning ‘waiting lists’ for elective surgery into ‘preparation lists’. British Journal of Anaesthesia, 126, 1–5.

Moyer, R., Ikert, K., Long, K. and Marsh, J. (2017). The Value of Preoperative Exercise and Education for Patients Undergoing Total Hip and Knee Arthroplasty: A Systematic Review and Meta-Analysis. JBJS Reviews, 5, e2.

Ng, S.X., Wang, W., Shen, Q., Toh, Z.A. and He, H.G. (2021). The effectiveness of preoperative education interventions on improving perioperative outcomes of adult patients undergoing cardiac surgery: a systematic review and meta-analysis. Eur J Cardiovasc Nurs.

Potts, G., Reid, D. and Larmer, P. (2022). The effectiveness of preoperative exercise programmes on quadriceps strength prior to and following anterior cruciate ligament (ACL) reconstruction: A systematic review. Physical Therapy in Sport, 54, 16–28.

Powell, R., Scott, N.W., Manyande, A., Bruce, J., Vögele, C., Byrne-Davis, L.M.T., Unsworth, M., Osmer, C. and Johnston, M. (2016). Psychological preparation and postoperative outcomes for adults undergoing surgery under general anaesthesia. Cochrane Database of Systematic Reviews.

Rodrigues, S.N., Henriques, H.R. and Henriques, M.A. (2021). Effectiveness of preoperative breathing exercise interventions in patients undergoing cardiac surgery: A systematic review. Revista Portuguesa de Cardiologia (English Edition), 40, 229–244.

Royal College Of Surgeons Of England (2020). Recovery of surgical services during and after COVID-19. Royal College of Surgeons of England. Available at: https://www.rcseng.ac.uk/coronavirus/recovery-of-surgical-services/

Royal College Of Surgeons Of England (2021). A new deal for surgery. Royal College of Surgeons of England. Available at: https://www.rcseng.ac.uk/-/media/7534--rcs--new-deal-for-surgery_aw3_web--270521.pdf

Shea, B.J., Reeves, B.C., Wells, G., Thuku, M., Hamel, C., Moran, J., Moher, D., Tugwell, P., Welch, V., Kristjansson, E. and Henry, D.A. (2017). AMSTAR 2: a critical appraisal tool for systematic reviews that include randomised or non-randomised studies of healthcare interventions, or both. BMJ, 358, j4008.

Thomsen, T., Villebro, N. and Møller, A.M. (2014). Interventions for preoperative smoking cessation. Cochrane Database of Systematic Reviews.

Tong, F., Dannaway, J., Enke, O. and Eslick, G. (2020). Effect of preoperative psychological interventions on elective orthopaedic surgery outcomes: a systematic review and meta-analysis. ANZ Journal of Surgery, 90, 230–236.

Van Der Gucht, E., Dams, L., Haenen, V., Godderis, L., Morlion, B., Bernar, K., Evenepoel, M., De Vrieze, T., Vandendriessche, T., Asnong, A., Geraerts, I., Devoogdt, N., De Groef, A. and Meeus, M. (2021). Effectiveness of perioperative pain science education on pain, psychological factors and physical functioning: A systematic review. Clinical Rehabilitation, 35, 1364–1382.

Wang, D., Wu, T., Li, Y., Jia, L., Ren, J. and Yang, L. (2021). A systematic review and meta-analysis of the effect of preoperative exercise intervention on rehabilitation after total knee arthroplasty. Ann Palliat Med, 10, 10986–10996.

Wang, L., Lee, M., Zhang, Z., Moodie, J., Cheng, D. and Martin, J. (2016). Does preoperative rehabilitation for patients planning to undergo joint replacement surgery improve outcomes? A systematic review and meta-analysis of randomised controlled trials. BMJ Open, 6, e009857.

Yau, D.K.W., Underwood, M. J., Joynt, G.M. and Lee, A. (2021). Effect of preparative rehabilitation on recovery after cardiac surgery: A systematic review. Annals of Physical and Rehabilitation Medicine, 64, 101391.

